# White matter hyperintensity spatial patterns: risk factors and clinical correlates

**DOI:** 10.1101/2024.06.27.24308274

**Authors:** Frauke Beyer, Ami Tsuchida, Aicha Soumaré, Hema Sekhar Reddy Rajula, Aniket Mishra, Fabrice Crivello, Markus Loeffler, Christophe Tzourio, Philippe Amouyel, Arno Villringer, Markus Scholz, Hélène Jacqmin-Gadda, Marc Joliot, A. Veronica Witte, Carole Dufouil, Stéphanie Debette

**Author notes:** **Corresponding author:** Stéphanie Debette, University of Bordeaux, INSERM, Bordeaux Population Health Center, UMR1219, 146 rue Léo Saignat, 33076 Bordeaux, France.

## Abstract

**INTRODUCTION:** White matter hyperintensities (WMH), a major cerebral small vessel disease marker, may arise from different pathologies depending on their location. We explored clinical and genetic correlates of agnostically derived spatial WMH patterns in two longitudinal population-based cohorts (3C-Dijon, LIFE-Adult).

**METHODS:** We derived seven WMH spatial patterns using Bullseye segmentation in 2,736 individuals aged 65+ and explored their associations with vascular and genetic risk factors, cognitive performance, dementia and stroke incidence.

**RESULTS:** WMH in the fronto-parietal and anterior periventricular region were associated with blood pressure traits, WMH genetic risk score (GRS), baseline and decline in general cognitive performance, incident all-cause dementia and ischemic stroke. Juxtacortical-deep occipital WMH were not associated with vascular risk factors and WMH GRS, but with incident all-cause dementia and intracerebral hemorrhage.

**CONCLUSION:** Accounting for WMH spatial distribution is key to decipher mechanisms underlying cerebral small vessel disease subtypes, an essential step towards personalized therapeutic approaches.

**Highlights:**

- We studied spatial patterns of white matter hyperintensities in 2736 participants
- Blood pressure was associated with fronto-parietal and anterior periventricular WMH
- Anterior periventricular WMH predicted dementia and stroke risk
- Juxtacortical-deep occipital WMH burden was not associated with blood pressure or WMH genetic risk
- Juxtacortical-deep occipital WMH predicted dementia and intracerebral hemorrhage.

**Research in context:**

1. **Systematic Review**: We performed a literature search using PubMed and Google Scholar. Most literature focuses on risk factors and outcomes for total WMH volume. Existing approaches to study WMH spatial patterns are heterogenous and often performed in single cohorts only.
2. **Interpretation**: Across two independent cohorts of older community-dwelling individuals, we show that WMH in the anterior periventricular and juxtacortical-deep occipital WMH show differential associations with risk factors and clinical outcomes. Anterior periventricular WMH were associated with blood pressure (BP) traits, WMH genetic risk, cognitive performance, dementia and incident stroke. Juxtacortical-deep occipital WMH did not relate to BP or WMH genetic risk and were predictive of dementia and intracerebral hemorrhage. These findings are important for characterizing small vessel disease subtypes and developing targeted prevention and therapeutic strategies.
3. **Future Directions**: Additional research is warranted to validate these findings and relate them to underlying vascular pathologies.

## 1 Introduction

Cerebral small vessel disease (cSVD) is a leading cause of ischemic and hemorrhagic stroke, cognitive decline and dementia [1] with no specific mechanism-based treatment to date. Covert cSVD, detectable on brain imaging in the absence of clinical disease, is highly prevalent in the general population with increasing age and portends a considerably increased risk of stroke, cognitive decline and dementia, thus representing a major target to prevent these disabling conditions and promote healthier brain aging [1,2]. However, guidelines on the use of available vascular prevention drugs for covert cSVD are based on limited evidence, with multiple situations of clinical equipoise [3]. This is at least partly related to the fact that cSVD is most often studied as a single entity, although its underlying biology and pathological substrate are highly heterogeneous, encompassing arteriolosclerosis, cerebral amyloid angiopathy (CAA), atherosclerosis at the origin of small perforating arteries [4], or features specific to rare inherited cSVD [5]. Molecular mechanisms of cSVD are poorly understood.

White matter hyperintensities of presumed vascular origin (WMH) represent the most common brain magnetic resonance imaging (MRI) marker of cSVD [6]. WMH frequently appear around the horns and borders of the lateral ventricles and in the centrum semiovale [7,8] but show considerable variability in spatial distribution [9,10]. This may reflect different underlying pathological processes and lead to distinct clinical manifestations [11,12]. CAA-associated WMH was for instance shown to have a multi-spot pattern in subcortical WM and a predominantly posterior distribution [13–15]. However, while extensive literature is available on determinants and consequences of global WMH burden, studies on determinants and clinical significance of spatial WMH patterns are scarce and heterogeneous.

Most studies on spatial WMH patterns have focused on a broad distinction between periventricular (PV) and deep WMH [16,17] while refined multivariate- and voxel-based approaches are less common [10]. Histopathological correlates across locations include gliosis, demyelination and axonal loss, the extent of which is associated with the severity of global WMH burden on brain MRI [18]. In PV WMH specifically, disruption of ependymal lining, increased extracellular water, myelin loss around tortuous venules and remyelination have been described [19–21] while in deep WMH, axonal loss, demyelination and hypoxia are more pronounced [22].

Besides age, high blood pressure (BP) is the most important risk factor for WMH [12,23,24], the strongest associations being described with PV and deep frontal WMH [8,17,25–27]. Higher body mass index (BMI) and waist-to-hip ratio (WHR) have been associated more prominently with deep WMH, possibly mediated by systemic inflammation [8,17,28]. Other vascular risk factors have not shown consistent relationships with spatial WMH patterns [10]. WMH is also a heritable trait with estimates ranging between 55 and 78% in family-based studies [29]. Large collaborative genome-wide association studies have identified common genetic variants in >25 genomic regions to be associated with larger total WMH volume [30]. However, apart from three loci specifically associated with PV WMH volume, little is known about genetic underpinnings of regional WMH patterns [23,31]. Some region-specific associations with WMH have been described for APOE-ε4, the most important genetic risk factor for Alzheimer’s disease (AD), with associations described to be more prominent with occipital, temporal [8] and parietal periventricular WMH [27]. APOE-ε2, protective for AD but a risk factor for CAA [32,33], has been associated with larger total WMH volume [34], but information on its association with WMH spatial patterns is lacking. In terms of clinical significance, PV WMH burden was most consistently associated with executive function, general cognitive decline and dementia risk [35–37]. Parietal or temporal WMH were suggested to specifically predict AD risk [38,39]. Overall, while some differential associations of spatial WMH patterns with risk factors and clinical outcomes have been described in population-based cohorts, a comprehensive assessment of these relationships for agnostically defined WMH spatial distributions is lacking. Here we derived WMH spatial patterns from a principal component analysis of WMH bullseye segmentation in two independent, large French and German population-based cohorts, and explored their association with vascular and genetic risk factors, cognitive performance and decline, and with risk of incident dementia and stroke.

## 2 Methods

### 2.1 Study populations

The Three-City Study (3C) is a longitudinal population-based cohort study comprising 9,294 participants, conducted in three French cities, Dijon, Bordeaux, and Montpellier, aiming to determine the risk of dementia attributable to vascular risk factors and disease [40] in non-institutionalized participants aged 65 years and older. The present analysis was conducted in the 3C-Dijon study (N=4,931), which has the largest brain MRI sub-study with standardized acquisitions performed at a single center on a 1.5T MRI scanner. High quality images at baseline were available in 1,670 participants without a history of stroke or dementia of which 1510 had genome-wide genotypes (**Supplementary Figure 9**). Sociodemographic and medical features along with cognitive performance were assessed at baseline and at follow-up examinations every 2-3 years over twelve years.

LIFE-Adult is a longitudinal population-based cohort study including 10,000 individuals from the Leipzig area aiming to study common diseases. In a subset of 2,765 participants, 3T MRI and cognitive testing were available [41,42]. The present study comprised 1,066 participants aged 65 years and older at baseline, without prevalent stroke or dementia, who had sufficient MRI quality and completed MRI preprocessing without errors based on visual inspection. Of these, 805 also had genome-wide genotype data available (**Supplementary Figure 10**).

Participants signed an informed consent and study protocols were approved by the ethics committees of Kremlin-Bicêtre University Hospital and Leipzig University. Both studies were conducted in agreement with the Declaration of Helsinki.

### 2.2 Vascular risk factors and other covariates

Hypertension (HTN) was defined as either having high BP (systolic BP (SBP) >140 mmHg and/or diastolic BP (DBP) >90 mmHg), a self-report of HTN, or use of antihypertensive drugs in both cohorts. In 3C-Dijon, BP was averaged over two measurements in a seated or lying and standing position. One participant with an outlying SBP value > mean + 5 standard deviations (SD) was excluded. In LIFE, BP was averaged over three measurements at 3-min intervals in a seated position. In both studies, weight, height, waist and hip circumferences were taken by trained study staff and used to calculate BMI as weight divided by height squared and WHR as waist divided by hip circumference. For analyses, we sex-standardized WHR using linear regression and applied the WHO WHR cutoff for abdominal adiposity defined as >0.9 for men and >0.85 for women to create a binary variable. Diabetes was defined as fasting glucose > 6.1 mmol/L or antidiabetic drug intake in 3C-Dijon, and based on self-reported diagnosis of diabetes, antidiabetic drug intake or HbA1c >6.5% in LIFE. Hypercholesterolemia was defined as total cholesterol >6.20 mmol/L and smoking status based on self-report in both studies: never, previous and current smoker. Education was defined in five categories in 3C-Dijon (no formal education, primary education with diploma, secondary education short, secondary education long and higher education) and in four categories in LIFE-Adult (no secondary school degree, secondary school degree, advanced secondary school degree and university entrance degree). In 3C-Dijon and LIFE-Adult, two and 27 participants were excluded because of missing education status (**Supplementary Tables 1-3**)

### 2.3 Genetic risk factors

In both studies, blood was collected at baseline and DNA was extracted from peripheral leukocytes. *APOE* genotyping was performed using PCR. In 3C-Dijon, genotyping was done on an Illumina Human 610-Quad BeadChip and imputed using MACH (version 1.0) and in LIFE, the Affymetrix Axiom CEU1 array and IMPUTE2 were used [40,41]. Both studies imputed, on the HRC and 1000 Genomes Phase 1 reference panel. We calculated a weighted genetic risk score (GRS) for total WMH volume in 3C-Dijon and LIFE based on 25 independent single nucleotide polymorphisms (SNPs) associated with WMH volume at genome-wide significance in the largest published European ancestry GWAS meta-analysis [23]. One of 25 SNPs was not available in 3C-Dijon (rs6797002) and a proxy (rs6809769) with r^2^ > 0.98 identified by LdProxy was used for the GRS and SNP analyses. We calculated GRS by summing the number of independent risk alleles associated at genome-wide significance (p<5.0×10^-8^) with WMH volume in the latest largest published GWAS [23]weighting each risk allele by the regression coefficient for the corresponding SNP in the published GWAS (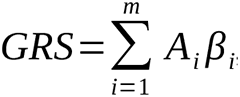, where *i* is a SNP among *i*m independent SNPs, *A* is the risk allele for SNP *i* and *β* is the regression coefficient in the published GWAS). Only SNPs with imputation score>0.9 and MAF>0.01 were included in GRS calculations.

### 2.4 Cognitive assessment

3C-Dijon participants underwent a neuropsychological test battery including the Isaacs Set Test (IST) [43], the Trail Making Test (TMT) [44], and the Benton Visual Retention Test (BVRT) [45], at baseline, after 2 years (except the TMT), 4, 8 and 12 years [46]. The IST is a verbal fluency test for which participants are prompted to generate as many words as possible in four semantic categories, within 60 seconds. The BVRT, a test of visual memory performance, consists of asking participants to recognize figures from a set of 15 stimuli among four options. The TMT assesses processing speed (part A), executive function and mental flexibility, by requesting participants to respectively connect numbers (part A) or numbers and letters in alternating order (part B) as quickly as possible [47,48].

For statistical analyses, we used the total number of words generated (IST), the total number of figures recognized (BVRT, range 0 to 15), and the time to complete part A and B with a cutoff at 300 seconds (TMT) (**Supplementary Figure 11**). LIFE participants underwent the Consortium to Establish a Registry for Alzheimer’s Disease (CERAD)-plus test-battery [49] which includes a phonemic and semantic fluency test, a word list memory task, and the TMT, at baseline and at 6.5 years follow-up. Verbal phonemic and semantic fluency was assessed by asking the participants to produce as many S-words and animal names as possible in 60 seconds. In the word list test, participants had to reproduce 10 items presented verbally over three trials and were asked to recall the 10 items after a delay and to recognize them in a list comprising 10 additional items not presented earlier. The TMT was performed as in 3C-Dijon. For statistical analyses, we used the total number of words produced in the phonemic and semantic verbal fluency tests, the raw scores of the word list learning (sum of all three rounds, ranging from 0-30), the raw scores of the word list delayed recall (ranging from 0-10), and the time to complete part A and B with a cutoff at 300 seconds.

### 2.5 Ascertainment of incident stroke and dementia

In 3C-Dijon, incident stroke and dementia were ascertained at each follow-up visit. A stroke was defined as a new focal neurological deficit of sudden or rapid onset, of presumed vascular origin, that lasted 24 hours or more, or leading to death [50]. If stroke symptoms or a hospitalization for stroke were reported, further information was collected from emergency medical service or hospitalization reports, neuroimaging reports, and interviews with the patient’s physician or the family [51]. The diagnosis of stroke and its type (ischemic or hemorrhagic) was confirmed by a panel of neurologists [51]. For the diagnosis of incident dementia, trained psychologists first screened participants with a detailed neuropsychological examination, and referred those participants who screened positive based on results of the Mini Mental State Examination and the IST to a neurologist. Cut-off scores were defined according to educational attainment [52]. A panel of neurologists took the final decision based on the Diagnostic and Statistical Manual of Mental Disorders, Fourth Revision (DSM-IV) criteria considering all information acquired in the study [40,53]. Probable and possible AD were diagnosed using the National Institute of Neurological and Communication Disorders and Stroke and of the Alzheimer’s Disease and Related Disorders Association criteria. We only included individuals without prevalent dementia and stroke, respectively. For LIFE, incident stroke and dementia cases were not available for analyses.

### 2.6 MRI Acquisition and pre-processing

In 3C-Dijon, anatomical T1-weighted, T2-weighted, and proton-density (PD)-weighted images were acquired on a 1.5 Tesla Siemens MAGNETOM scanner [40]. T1-weighted images were acquired using a 3D inversion recovery fast spoiled-gradient echo sequence (TR = 97 ms, TE = 4 ms, TI = 600 ms, voxel size = 1.0×0.98×0.98 mm^3^) and T2- and PD-weighted images using a 2D fast spin echo sequence with two echo times (TR = 4400 ms, TE1 = 16 ms, TE2 = 98 ms). T2 and PD acquisitions had 35 axial slices with 3.5 mm slice thickness and 0.5 mm spacing between slices, with an in-plane resolution of 0.98×0.98 mm^2^. A multispectral segmentation was applied to first detect WM voxels and then determine low- and high-contrast WMH voxels [54]. We used the full WMH probability maps including both low and high-contrast WMH voxels. T1-weighted imaging was processed using FreeSurfer version 7.3.2 to derive anatomical labels used for Bullseye segmentation (see 2.7 Bullseye WM segmentation) and total intracranial volume (TIV).

In LIFE, anatomical T1-weighted and fluid-attenuated inversion-recovery (FLAIR) images were acquired on a 3 Tesla Siemens MAGNETOM Verio scanner with a 32-channel head coil. T1-weighted imaging used a rapid acquisition gradient echo sequence (TR = 2,300 ms, TI = 900 ms, TE = 2.98 ms, voxel size = 1 mm^3^), with different parameters used for FLAIR images (TR = 5,000 ms, TI = 1,800 ms, TE = 395 ms, voxel size = 1×1×1 mm^3^). The cross-sectional pipeline of the Lesion Segmentation Toolbox (version 3.0.0, run on MATLAB version 9.10) with the Lesion Prediction algorithm and default parameters was used to estimate WMH probability maps [55]. T1-weighted imaging was processed with FreeSurfer version 5.3.0 to derive individual anatomical labels and TIV [56].

### 2.7 Bullseye WMH segmentation

We extracted regional WMH volumes based on the previously described Bullseye WM segmentation in each subject’s native space [57]. Briefly, FreeSurfer output was used to generate a WM segmentation according to normalized distance between ventricles and cortex (4 shells) and approximate lobar/brain region (frontal, temporal, parietal, occipital on both hemispheres, basal ganglia, **Figure 1A**) [58]. For each participant this segmentation was affine-registered to the FLAIR/T2-weighted image using FSL’s FLIRT for 3C-Dijon and FreeSurfer’s bbregister algorithms for LIFE due to issues with the FreeSurfer processing in 3C-Dijon. WMH volumes were extracted as the sum of WMH probabilities in each of the 36 brain regions (frontal, temporal, parietal, occipital on both hemispheres + basal ganglia in four shells [57]). WMH volumes in the Bullseye parcels were scaled by multiplying with the ratio of individual to average TIV in the respective cohort [59].

**Figure 1:**
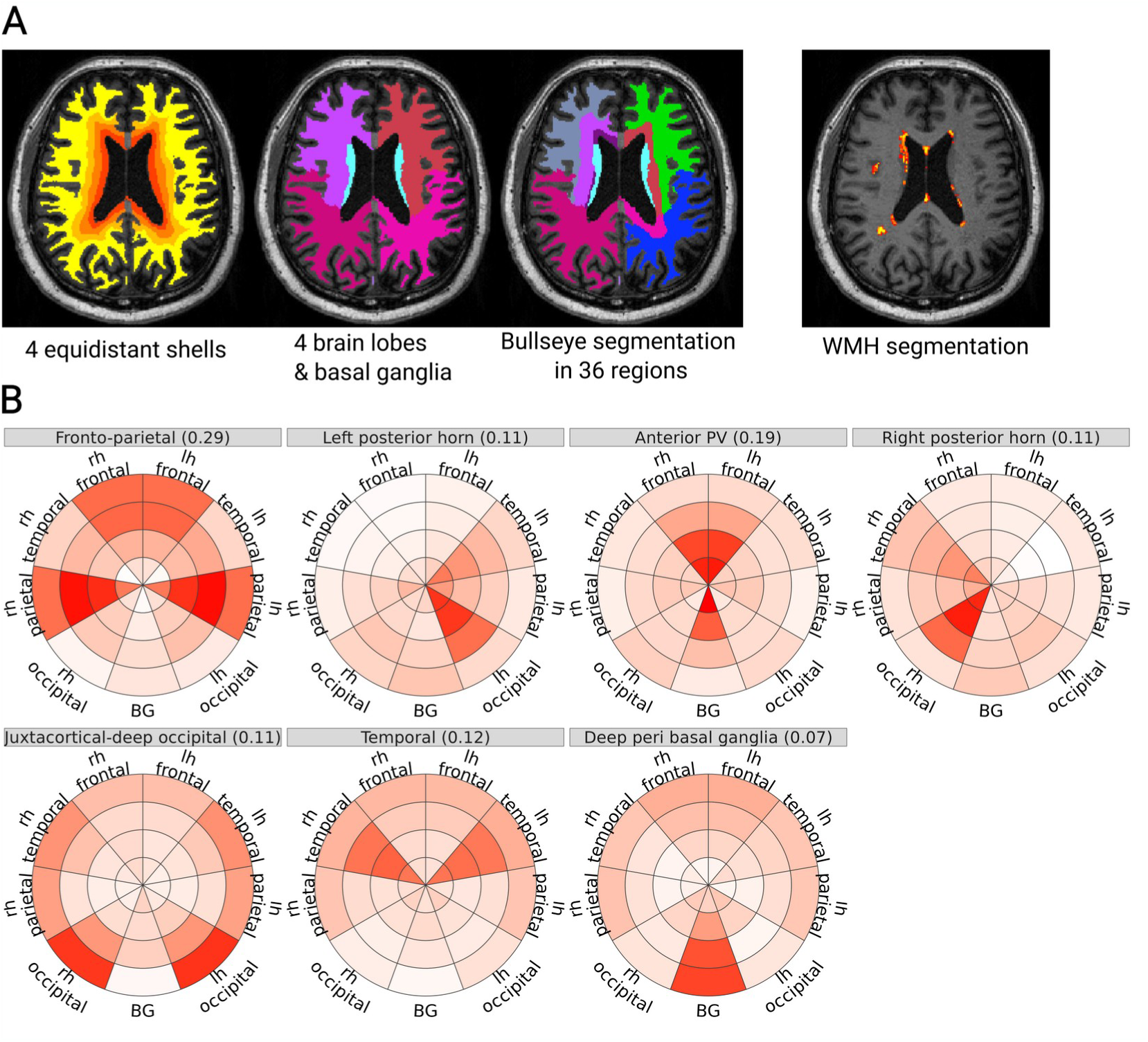
Bullseye Segmentation and WMH spatial components. **A**: Illustration of the Bullseye approach for WMH spatial segmentation: left: four equidistant shells between ventricles and cortex, middle left: WM is classified as belonging to four brain lobes on each hemisphere or basal ganglia region, middle right: combination of shells and lobar segmentation yields Bullseye segmentation with 36 WM regions, right: WMH probability map which is summed in each region to the spatial WMH volume. **B**: WMH spatial components derived from PCA in the 3C cohort. Label indicates name and variance explained in brackets, color indicates loading of the region, distance from center represents four equidistant shells between ventricles and cortex, and angular orientation represents cerebral lobes and hemisphere. WMH: white matter hyperintensities, WM: white matter, PCA: principal component analysis, TC: transformed components.

### 2.8 Principal Component Analysis

We used principal component analysis (PCA) to extract WMH spatial components [58]. Given the larger sample size, we performed the initial PCA in 3C-Dijon and applied the decomposition to LIFE-Adult. We used parallel analysis implemented in `fa.parallel` to determine the optimal number of components and applied `oblimin` rotation under the assumption that WMH spatial components are not independent and for better interpretability (**Supplementary Figure 4**). Components derived from a PCA generated in LIFE-Adult (sensitivity analysis) corresponded closely to those derived from 3C-Dijon (**Supplementary Figures 1-3**). Component scores for each individual from 3C-Dijon resulted from the PCA, and component scores for individuals from LIFE-Adult were derived by projecting the data onto the 3C-Dijon PCA components.

### 2.9 Statistical Analysis

All analyses were performed in R version 4.2.2.

#### 2.9.1 Association of WMH spatial components with genetic and vascular risk factors

We tested two sets of models: Our primary analyses used linear regression with individual risk factors (HTN, SBP, DBP, WHR (sex-standardized continuous and binarized), BMI, diabetes, hypercholesterolemia, smoking status (never, previous, current smoker), APOE-ε4 and APOE-ε2 carrier status) as predictors and WMH spatial components scores as outcomes, adjusting for age, sex and TIV. In secondary analyses we included multiple putative risk factors in the same linear regression model: SBP, DBP, BMI, hypercholesterolemia, diabetes, smoking and APOE-ε4 carrier status. We additionally explored differences between APOE -ε4 and ε2 homozygotes and either non-carriers or ε3 homozygotes as control groups. We tested the association of a GRS and of the 25 individual WMH risk SNPs with WMH spatial component scores in additive linear regression models, adjusted for age, sex, TIV and the first three components of population stratification. All these analyses were conducted separately in 3C-Dijon and LIFE-Adult, followed by inverse-variance weighted fixed effects meta-analyses using metagen [60]. For vascular risk factors, we meta-analyzed the coefficients of the primary and secondary linear models. P-values were adjusted for multiple comparisons using Benjamini-Hochberg false discovery rate correction (FDR) with pFDR < 5% for inference [61]. 120 tests were included for vascular/genetic analyses, 64 cognitive and 40 for dementia/stroke analyses. For comparison, we also assessed the relation of vascular risk factors and GRS with total WMH volume.

#### 2.9.2 Association of WMH spatial components with baseline and change in cognitive performance

We estimated the relationship between WMH spatial components and cognitive function (general cognitive function, memory and executive function) using latent class mixed models (LCMM) from the `lcmm` package [62]. LCMM combines latent variable and mixed modeling to estimate the effect of predictors on latent constructs. It takes into account the curvilinearity of psychometric tests and is therefore considered to be more accurate [63]. We assumed that latent processes underlie the respective scores for the aforementioned neuropsychological tests. For general cognitive function, we used TMT part A, TMT part B, BVRT and IST as indicators in 3C-Dijon and TMT-B, TMT-A, CERAD word list learning and recall scores, phonemic and semantic fluency in LIFE-Adult. In both studies, TMT-B was the indicator of executive function, while BVRT and CERAD word list learning and recall scores were used as respective indicators of visual and verbal memory in 3C-Dijon and LIFE-Adult. In 3C-Dijon, we compared base models which included age and sex as predictors with time as linear and linear + quadratic predictor of cognitive trajectories for each latent variable and selected the model with lowest Akaike information criterion (AIC). For executive and general cognitive function, linear effect of time yielded lower AIC while for memory, a model including both linear and quadratic terms was slightly superior. In LIFE-Adult, time was modeled as linear. We used an indicator for first assessment time point to adjust for practice effects in 3C-Dijon, and also adjusted for age at baseline, sex and education. In both cohorts, we used time since first assessment as time scale and splines with 3 nodes as non-linear link function.

We tested the association of all baseline WMH spatial components with the latent cognitive variables as well as their interaction with time, using FDR for multiple testing correction, and meta-analyzed results for associations of WMH spatial components with general cognitive and executive function using inverse-variance weighted meta-analysis. We also assessed the relation of total WMH volume with cognitive outcomes.

#### 2.9.3 Association of WMH spatial components with incident stroke and dementia

In 3C-Dijon, we analyzed the association of WMH spatial components with incident all-cause dementia and AD using Cox proportional hazards regression models with time-to-event since baseline as a time scale. We adjusted for age at baseline, sex, education level and TIV. We also investigated the association of WMH spatial components with risk of incident any stroke, ischemic stroke and intracerebral hemorrhage using Cox proportional hazards regression models adjusting for age at baseline, sex and TIV. We accounted for multiple testing across dementia and stroke subtypes separately using FDR-correction. Again, for comparison, we also assessed the relation of total WMH volume with dementia and stroke incidence.

## 3 Results

Our study sample comprised 2,736 older participants from the general population: 1,066 participants from the German LIFE-Adult study (mean age [SD], 70.5 [3.7] years; 489 [46%] women; mean follow-up [SD], 6.6 [0.6] years) and 1,670 participants from the French 3C-Dijon study (72.4 [4.1] years; 1,012 [60.6%] women, 5.6 [3.2] years follow-up). In 3C-Dijon, seven WMH spatial components were derived from the principal component analysis of the WMH Bullseye segmentation (**Figure 1B**) and corresponded well to components derived from LIFE-Adult (**Supplementary Figures 1-3**). These spatial patterns were characterized as fronto-parietal (component 1, C1), right posterior horn (C2), anterior PV (C3), left posterior horn (C4), juxtacortical-deep occipital (C5), temporal (C6), deep peri basal ganglia (C7).In 3C-Dijon, only fronto-parietal (C1) and anterior PV (C3) WMH were associated with age. In LIFE-Adult all components showed positive associations with age, with age explaining least variance for C5 (<1%) and most for C1 (10%) (**Supplementary Figures 5-8**).

### 3.1 Association of putative risk factors with WMH spatial patterns

Among vascular risk factors, expectedly, the strongest associations with WMH spatial components were observed for BP traits. In both cohorts, BP-related measures (DBP and to a lesser extent SBP) were significantly associated with fronto-parietal (C1) and anterior PV (C3) WMH, even after accounting for other vascular risk factors (**Figure 2**). The strongest associations were observed with anterior PV (C3) WMH for both SBP (meta-pFDR=8.8×10⁻^9^) and DBP (meta-pFDR=7.9×10⁻^12^). Higher WHR was also associated with greater scores of anterior PV (C3) WMH in multivariable models (meta-pFDR=0.032, meta-q=0.014), while higher BMI was associated with higher scores in temporal (C6) WMH (meta-pFDR=6.1×10⁻⁴, meta-q=2.6×10⁻⁴) and deep peri basal ganglia (C7) WMH (meta-pFDR=3.2×10⁻⁵, meta-q=1.2×10⁻^5^, **Table 2, Supplementary Table 4**). Meta-analyses showed no significant heterogeneity (I²<50%) except for SBP associations with fronto-parietal (C1) (I²=94%, random effects meta-p=0.22) and anterior PV (C3) WMH (I²=80%, random effects meta-p=0.001), DBP association with right posterior horn (C2) (I²=74%, random effects meta-p=0.096 and the association of BMI with temporal (C6) WMH (I²=74%, random effects meta-p=0.012).

**Figure 2:**
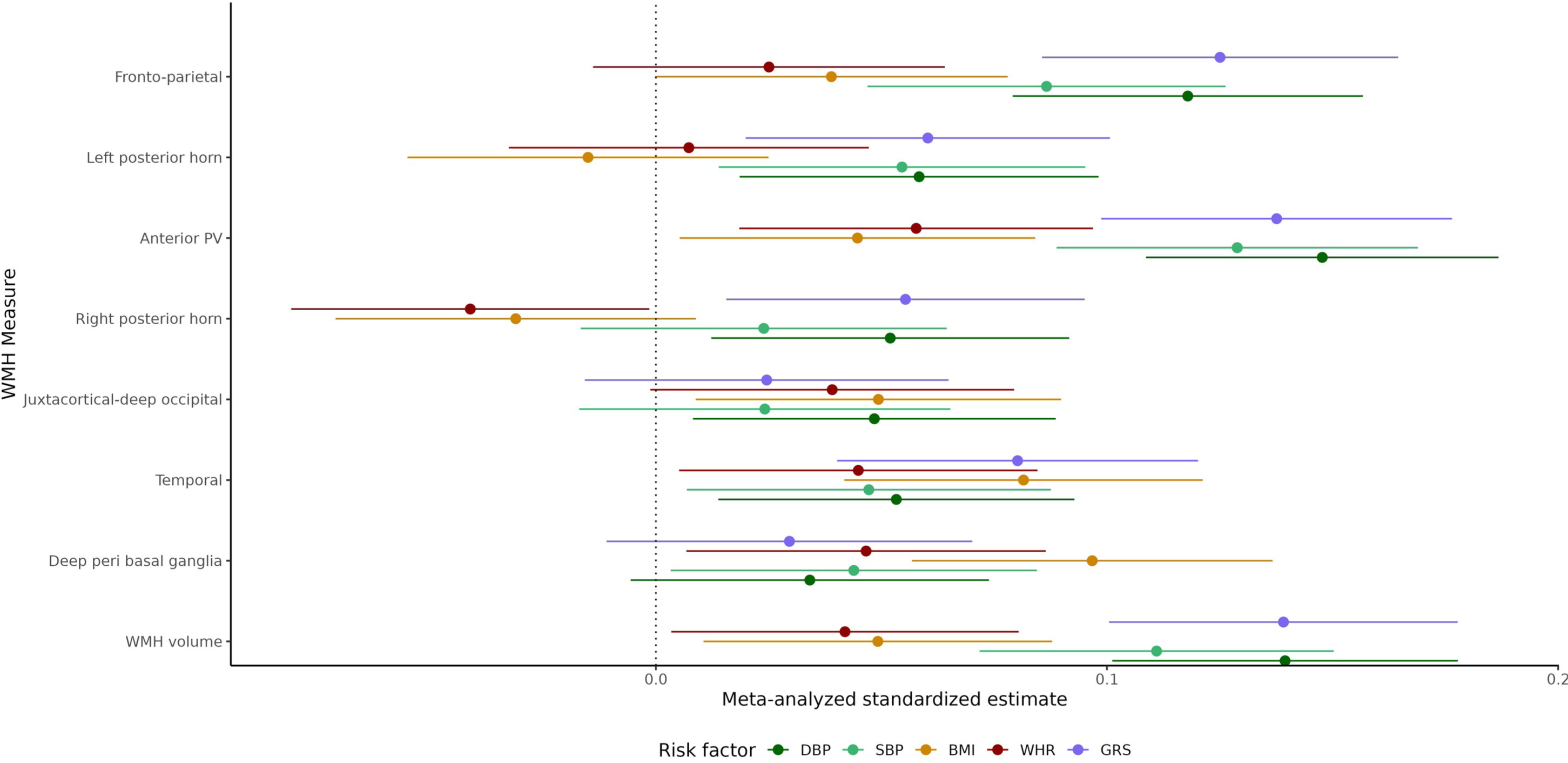
Associations of vascular risk factors and total WMH genetic risk score with WMH spatial components (meta-analysis of 3C-Dijon and LIFE-Adult) Forest plot showing the standardized regression estimates and 95% CI of DBP, SBP, BMI, WHR and GRS with WMH spatial components. All VR models adjusted for age, sex, education, TIV, DBP/BMI, hypercholesterolemia, smoking status, diabetes and APOE ε4 carrier status. All models for GRS adjusted for age, sex, TIV and 3 PC. SBP: systolic blood pressure, DBP: diastolic blood pressure, BMI: body mass index, WHR: waist-to-hip ratio, GRS: genetic risk score, CI: confidence interval, WMH: white matter hyperintensities, TIV: total intracranial volume, PC: principal component of population stratification.

**Table 1:** Demographic characteristics of 3C-Dijon and LIFE-Adult cohorts.

|  | 3C-Dijon |  |  | LIFE-Adult |  |  |
| --- | --- | --- | --- | --- | --- | --- |
|  | N | Mean/% | SD | N | Mean/% | SD |
| Age (y) | 1670 | 72.4 | 4.1 | 1066 | 70.5 | 3.65 |
| Females | 1012 | 60.6 |  | 577 | 54.1 |  |
| SBP (mmHg) | 1670 | 149 | 22.3 | 1025 | 134 | 16.6 |
| DBP (mmHg) | 1670 | 84.9 | 11.3 | 1025 | 74 | 9 |
| Hypertension (yes/%) | 1287 | 77.1 |  | 714 | 67% |  |
| Antihypertensives (yes/NA) | 704 | 42.2% |  | 660/7 | 62.3/1 |  |
| Smoking (never/previous/current/NA) | 1027/550/93 | 62/33/6 |  | 602/324/68/7<br>2 | 56/30/7/7 |  |
| Hypercholesterolemia | 1173/483/14 | 70/29/1 |  | 553/430/83 | 52/40/8 |  |
| Diabetes (yes/NA) | 183/16 | 11/1 |  | 211 | 20 |  |
| BMI (kg/m<sup>2</sup>) | 1669 | 25.4 | 3.74 | 1065 | 27.6 | 4.1 |
| WHR | 1648 | 0.869 | 0.085<br>2 | 1061 | 0.955 | 0.0834 |
| WHR binary (yes/NA) | 191/22 | 47/1 |  | 917 | 86 |  |
| ApoE ε4 (0/1/2/NA risk alleles) | 1286/350/21/13 | 77/21/1.3/1 |  | 756/229/16/6<br>5 | 71/21/1.5/<br>6 |  |
| ApoE ε2 (0/1/2/NA risk alleles) | 1657 | 85/13/0.5/0.<br>7 |  | 1001 | 81/12/0.7/<br>6 |  |

SBP: systolic blood pressure, DBP: diastolic blood pressure, BMI: body mass index, WHR: sex-standardized waist-to-hip ratio, ApoE: apolipoprotein

**Table 2:** Meta-analysis of VR factors and WMH spatial components in 3C-Dijon and LIFE-Adult.

| WMH Measure | VRF | 3C-Dijon |  |  |  |  | LIFE-Adult |  |  |  |  | Meta-analysis |  |  |  |
| --- | --- | --- | --- | --- | --- | --- | --- | --- | --- | --- | --- | --- | --- | --- | --- |
| | | $\beta$ | Adj. $\beta$ | SE | p | pFDR | $\beta$ | Adj. $\beta$ | SE | p | pFDR | $\beta$ | SE | p | pFDR |
| Fronto-parietal | SBP | 0.0224 | 0.021 | 0.026 | 0.42 | 0.69 | 0.171 | 0.197 | 0.033 | 4.05e-09 | 4.15e-07 | 0.087 | 0.020 | 1.89e-05 | 1.94e-04 |
| Fronto-parietal | DBP | 0.0963 | 0.097 | 0.025 | 1.07e-04 | 0.0012 | 0.157 | 0.155 | 0.033 | 2.65e-06 | 5.44e-05 | 0.118 | 0.020 | 2.60e-09 | 4.69e-08 |
| Left posterior horn | DBP | 0.0204 | 0.029 | 0.025 | 0.25 | 0.51 | 0.103 | 0.109 | 0.034 | 0.0012 | 0.011 | 0.058 | 0.020 | 0.004 | 0.029 |
| Anterior PV | SBP | 0.1088 | 0.099 | 0.025 | 9.59e-05 | 0.0012 | 0.177 | 0.184 | 0.034 | 1.25e-07 | 4.50e-06 | 0.129 | 0.020 | 2.79e-10 | 6.70e-09 |
| Anterior PV | DBP | 0.1486 | 0.143 | 0.025 | 8.64e-09 | 4.2e-07 | 0.154 | 0.157 | 0.034 | 3.94e-06 | 7.08e-05 | 0.148 | 0.020 | 1.25e-13 | 9.00e-12 |
| Anterior PV | WHR | 0.0610 | 0.041 | 0.025 | 0.094 | 0.3 | 0.094 | 0.091 | 0.035 | 0.0088 | 0.054 | 0.058 | 0.020 | 0.0039 | 0.029 |
| Temporal | BMI | 0.0447 | 0.047 | 0.025 | 0.063 | 0.24 | 0.110 | 0.142 | 0.034 | 2.85e-05 | 4.57e-04 | 0.082 | 0.020 | 5.83e-05 | 5.25e-04 |
| Deep peri basal ganglia | BMI | 0.1039 | 0.089 | 0.025 | 4.16e-04 | 0.004 | 0.075 | 0.112 | 0.035 | 0.0014 | 0.012 | 0.097 | 0.020 | 2.09e-06 | 2.51e-05 |
| WMH volume | SBP | 0.0711 | 0.064 | 0.025 | 0.01 | 0.059 | 0.183 | 0.194 | 0.033 | 8.30e-09 | 4.15e-07 | 0.111 | 0.020 | 2.94e-08 | 4.23e-07 |
| WMH volume | DBP | 0.1326 | 0.127 | 0.024 | 1.77e-07 | 5.1e-06 | 0.161 | 0.162 | 0.033 | 1.08e-06 | 2.59e-05 | 0.139 | 0.020 | 9.49e-13 | 3.42e-11 |

Shown are all associations significant in meta-analysis of fully adjusted models. For 3C-Dijon and LIFE-Adult, $\beta$ represents standardized regression estimates from linear regression models adjusting for age, sex and TIV; adjusted $\beta$ , standard error and p represent linear regression models fully adjusted for SBP or BMI, hypercholesterolemia, diabetes, smoking and APOE ε4 carrier status. For the IVW meta-analysis estimate, SE, p-value and FDR-p value are shown. SBP: systolic blood pressure, DBP: diastolic blood pressure, WHR: sex-standardized waist-to-hip ratio, WMH: white matter hyperintensities, FDR: false-discovery rate, SE: standard error, IVW: inverse-variance weighted; VRF: vascular risk factor

Regarding genetic risk factors, the weighted GRS for total WMH volume derived from the largest WMH GWAS meta-analysis was most strongly associated with fronto-parietal (C1), anterior (C3) and temporal (C6) WMH (meta-pFDR<0.001), the most significant association being observed, as for BP traits, with C3 WMH (meta-pFDR=2.5×10 ^-12^). However, heterogeneity was also highest for this component (I²=85%, random effects meta-p=0.01). The WMH GRS further showed significant association with right and left posterior horn (C2, C4) WMH (meta-pFDR<0.05), but no association with juxtacortical-deep occipital and deep peri basal ganglia (C5, C7) WMH, consistently in both studies (I²<50%, **Table 3**). In secondary analyses we explored individual associations of the 25 independent genetic risk variants included in the WMH GRS with WMH spatial components. The most significant risk variant for total WMH volume, rs34974290 at *TRIM47* (chr17q25), was associated with fronto-parietal (C1) WMH (meta-estimate ± standard error: 0.142±0.036, meta-pFDR=0.018) and anterior PV (C3) WMH (0.128±0.035, meta-pFDR=0.049). Neither APOE-ε4 nor APOE-ε2 carrier status (heterozygote or homozygote) were significantly associated with spatial WMH components (**Supplementary Table 5**).

**Table 3:** Associations of GRS with WMH spatial components in the 3C-Dijon and LIFE-Adult cohorts.

|  | 3C-Dijon |  |  |  | LIFE-Adult |  |  |  | Meta-analysis |  |  |  |
| --- | --- | --- | --- | --- | --- | --- | --- | --- | --- | --- | --- | --- |
| WMH Measure | $\beta$ | SE | p | pFDR | $\beta$ | SE | p | pFDR | $\beta$ | SE | p | pFDR |
| Fronto-parietal | 0.12 | 0.03 | 3.07e-06 | 1.64e-05 | 0.14 | 0.03 | 4.26e-05 | 4.26e-05 | 0.13 | 0.02 | 5.23e-10 | 1.39e-09 |
| Left posterior horn | 0.03 | 0.03 | 0.22 | 0.27 | 0.11 | 0.03 | 0.0013 | 0.0013 | 0.06 | 0.02 | 0.0034 | 0.0055 |
| Anterior PV | 0.17 | 0.02 | 8.64e-12 | 1.38e-10 | 0.08 | 0.03 | 0.019 | 0.019 | 0.14 | 0.02 | 3.98e-12 | 1.59e-11 |
| Right posterior horn | 0.04 | 0.03 | 0.14 | 0.19 | 0.09 | 0.03 | 0.0098 | 0.0098 | 0.06 | 0.02 | 0.0063 | 0.0084 |
| Juxtacortical-deep occipital | 0.03 | 0.03 | 0.27 | 0.31 | 0.02 | 0.04 | 0.61 | 0.61 | 0.02 | 0.02 | 0.23 | 0.23 |
| Temporal | 0.07 | 0.03 | 0.0091 | 0.017 | 0.10 | 0.03 | 0.0022 | 0.0022 | 0.08 | 0.02 | 8.54e-05 | 1.71e-04 |
| Deep peri basal ganglia | 0.01 | 0.03 | 0.56 | 0.6 | 0.06 | 0.04 | 0.093 | 0.093 | 0.03 | 0.02 | 0.15 | 0.17 |
| WMH volume | 0.15 | 0.02 | 6.91e-10 | 5.53e-09 | 0.12 | 0.03 | 5.53e-04 | 0.00055 | 0.14 | 0.02 | 1.68e-12 | 1.34e-11 |

Shown are standardized regression estimates $\beta$ from regression models adjusting for age, sex, 3 PC and TIV, FDR-corrected p-values for both cohorts and $\beta$ estimate, p-value and FDR-p from the IVW meta-analysis. . GRS: genetic risk score, WMH: white matter hyperintensities, FDR: false-discovery rate, SE: standard error, IVW: inverse-variance weighted, PC: principal components of population stratification, PV: periventricular.

### 3.2 Association of WMH spatial patterns with baseline and longitudinal change in cognitive performance

We investigated the association of the WMH components with baseline and longitudinal change in general cognitive function in 3C-Dijon and LIFE-Adult using latent process mixed models. Across cohorts, larger fronto-parietal (C1) and anterior PV (C3) WMH burden was consistently associated with worse baseline general cognitive function, and, especially C3, with more marked decline in general cognitive performance (meta-pFDR<0.05, I²<50%, **Table 4**). Total WMH volume was strongly related to baseline and change in general cognitive function in both cohorts and in the meta-analysis. Next, we explored the relation of WMH components with individual cognitive domains. Larger WMH burden in the frontal-parietal deep and anterior PV region (C1, C3) was associated with worse executive function at baseline (C1: meta-analysis estimate (ß) ± standard error (SE)=-0.13±0.03, meta-pFDR=2.4×10⁻^9^, I²<6%; C3: ß±SE=-0.11±0.03, meta-pFDR=3.3×10⁻^4^, I²=0%, **Supplementary Table 6**). Temporal and deep peri basal ganglia WMH (C6, C7) were also associated with worse executive function at baseline (C6: ß±SE=-0.07±0.03, meta-pFDR=0.014, I²=88%; C7: ß±SE=-0.1±0.03, meta-pFDR=8.04×10⁻^4^, I²=55%), but with high heterogeneity across cohorts, and in a random effects meta-analysis only the association with C7 remained significant (meta-p=0.01). Total WMH volume was also significantly associated with worse executive function at baseline (ß±SE=-0.13±0.03, meta-pFDR=1.18×10⁻^5^, I²=0). We observed no association of WMH spatial components or total volume with change in executive function over time. 3C-Dijon and LIFE-Adult tested different aspects of memory, i.e. visual and verbal memory respectively, which could not be meta-analyzed. In 3C-Dijon, WMH spatial components and total WMH volume did not show any significant association with baseline or change (interaction with linear or quadratic time) in visual memory after multiple testing correction (**Supplementary Table 7**). In LIFE-Adult, fronto-parietal (C1) and left posterior horn (C2) WMH burden was associated with worse baseline verbal memory performance (standardized estimate ±SE: -0.16±0.04, pFDR=0.004 and - 0.14±0.04, pFDR=0.03 for C1 and C2), but not with change in memory performance (**Supplementary Table 8**).

**Table 4:**
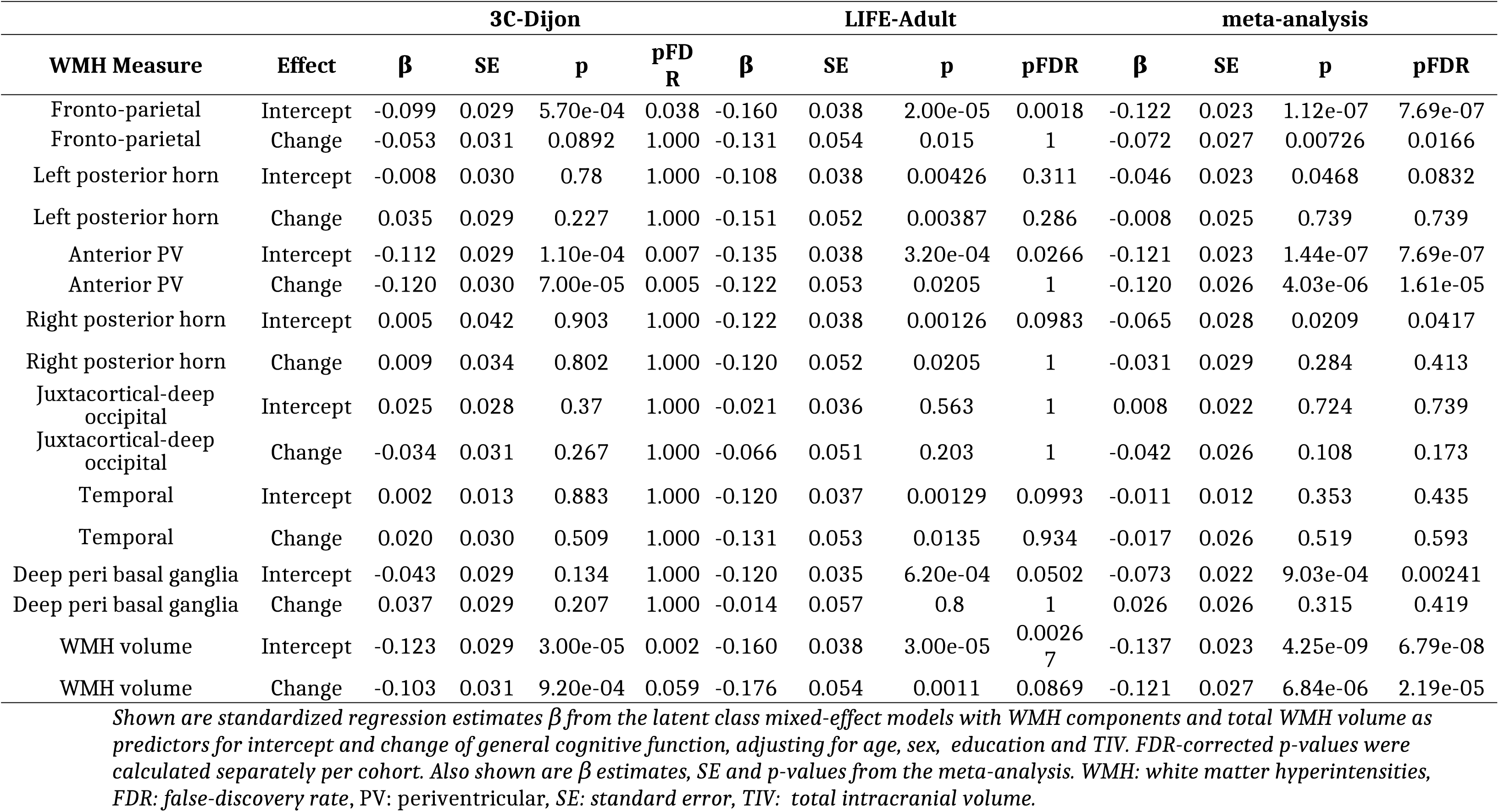
Associations of WMH spatial components with baseline and change in general cognitive function in 3C-Dijon and LIFE-Adult.

### 3.3 Associations of WMH spatial patterns with incident dementia and stroke

Information on the occurrence of incident dementia and stroke was available in 3C-Dijon only. Over up to 12 years, out of 1,668/1,644 participants with follow-up and no prevalent dementia or stroke at baseline, 118 developed incident all-cause dementia (of whom 87 probable or possible AD) and 64 suffered an incident stroke (50 ischemic strokes, 10 intracerebral hemorrhages and 4 strokes of unknown type). Anterior PV (C3) and juxtacortical-deep occipital (C5) WMH were significantly associated with a higher risk of incident all-cause dementia (Hazard Ratio [95% confidence interval], HR[95%CI]= 1.27[1.05-1.53], p=0.012, pFDR=0.08 and 1.22[1.005 -1.45], p=0.045, pFDR=0.27). Participants in the highest quartile of anterior PV (C3) and juxtacortical-deep occipital (C5) WMH distribution had a roughly two-fold higher risk of dementia (HR[95%CI]=2.39[1.28,4.44], p=0.0058 and HR[95%CI]=1.87[1.14,3.08], p=0.014, Figure 4 and **Supplementary Table 9**). None of the WMH components was significantly associated with AD (**Supplementary Table 10**).

**Figure 4:**
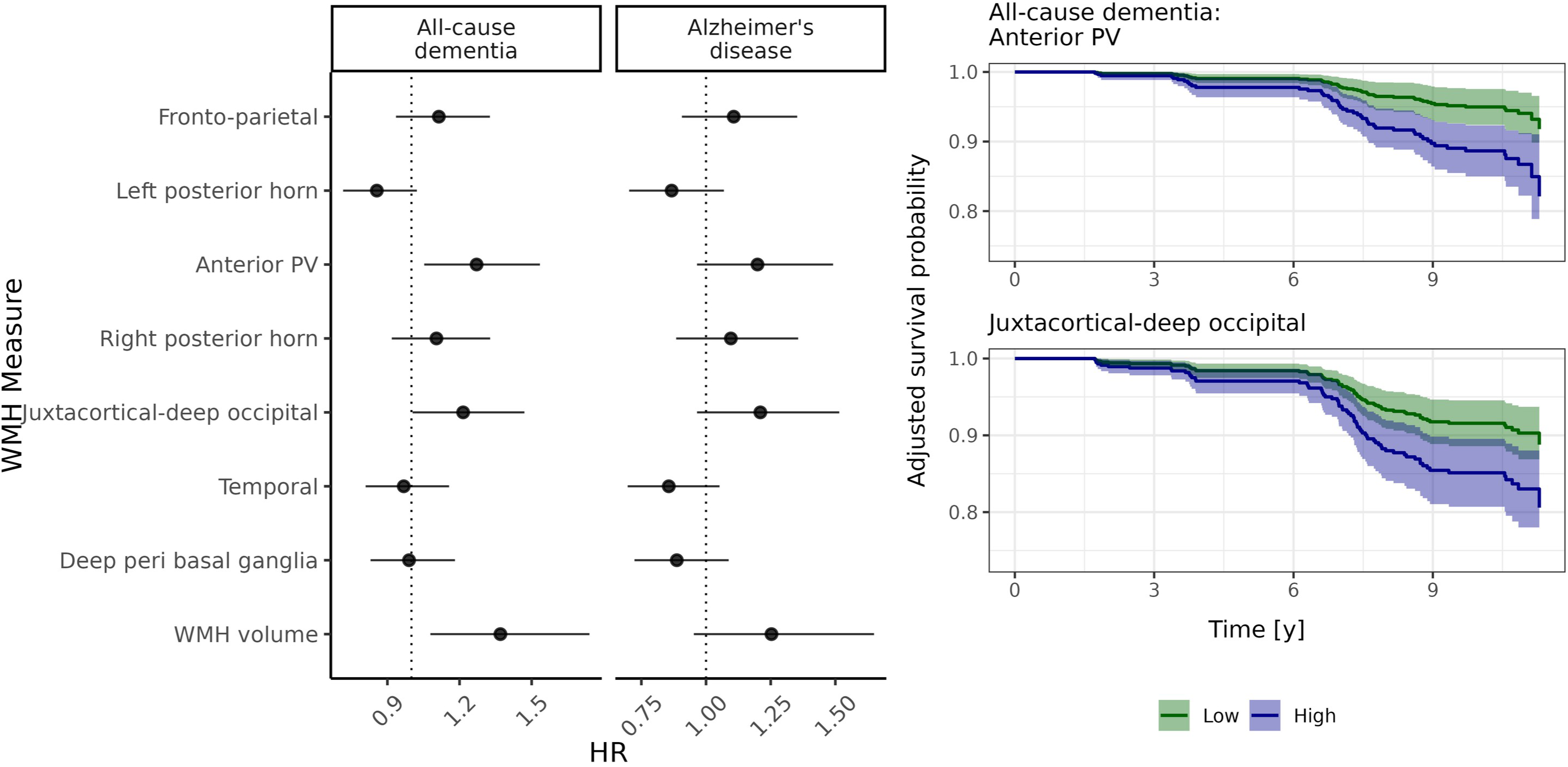
WMH spatial components and incident all-cause dementia and Alzheimer’s disease in 3C-Dijon. Left: Forest plot shows HR and 95% CI of the Cox survival model for all-cause and probable or possible dementia for each component adjusting for age, sex, education and TIV. Right: Adjusted survival probabilities for all-cause dementia from WMH component models with 95% CI. Shown are curves for highest and lowest quartile of WMH components 3 and 5. HR: hazard ratio, CI: confidence interval, WMH: white matter hyperintensities, TIV: total intracranial volume.

Fronto-parietal (C1), anterior PV (C3) and juxtacortical-deep occipital (C5) WMH were all associated with increased risk of incident any stroke (HR[95%CI]=1.39[1.09-1.78], p=0.0073, pFDR=0.12, q=0.025; HR[95%CI]=1.67[1.28-2.18], p=0.00015, pFDR=0.036; HR[95%CI]=1.32[1.02-1.72], p=0.037, pFDR=0.56 **Supplementary Table 11**). Participants in the highest quartile of anterior PV (C3) WMH distribution had a nearly four-fold risk of developing any stroke (HR[95%CI]=3.88 [1.75-8.63], p=8.7×10^-4^, while participants in the highest quartile of fronto-parietal (C1) WMH had a nearly three-fold risk (HR[95%CI]=2.94 [1.39-6.21], p=0.0047, Figure 5).

**Figure 5:**
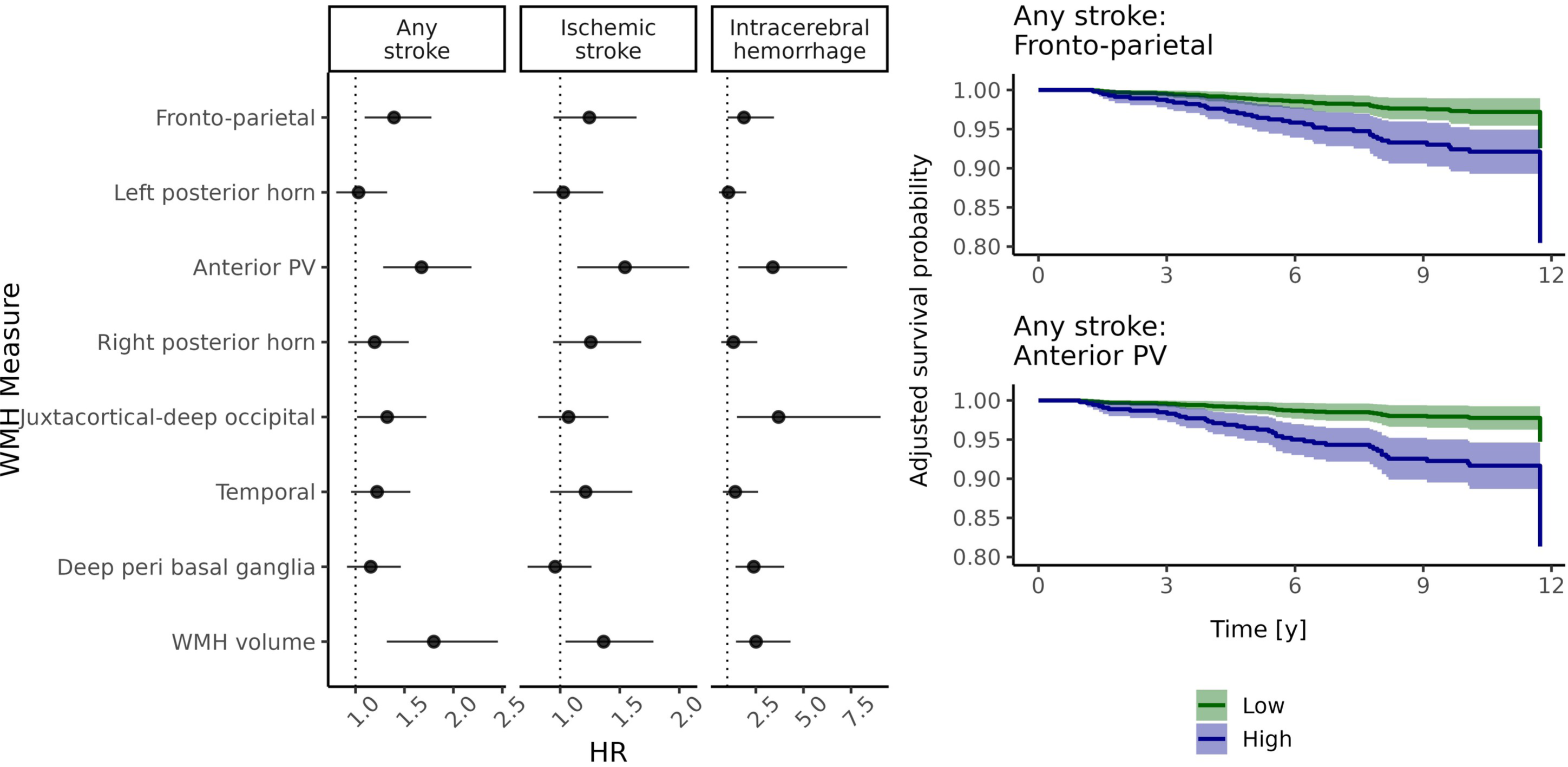
WMH spatial components and incident stroke in 3C-Dijon. Left: Forest plot shows HR and 95% CI of the Cox survival model for any stroke, ischemic stroke and intracerebral hemorraghe for each component adjusting for age, sex, education and TIV. Right: Adjusted survival probabilities for any stroke with 95% CI. Shown are curves for highest and lowest quartile of WMH component 3. HR: hazard ratio, CI: confidence interval, WMH: white matter hyperintensities, TIV: total intracranial volume.

Anterior PV (C3) WMH were associated with both incident ischemic stroke (HR[95%CI]=[1.14-2.08], p=0.004, pFDR=0.076) and intracerebral hemorrhage (HR[95%CI]=3.39[1.58-7.29], p=0.002, pFDR=0.04). Higher scores of juxtacortical-deep occipital (C5) and deep basal ganglia (C7) WMH were associated with increased risk of intracerebral hemorrhage (HR[95%CI]=3.69[1.51-9.05], p=0.004, pFDR=0.076; HR[95%CI]=2.39[1.43-3.99], p=0.001, pFDR=0.021, **Supplementary Tables 12-13**).

## 4 Discussion

Using the Bullseye method for white matter parcellation, we derived seven spatial components of WMH in two independent older population-based cohort studies comprising 2,736 participants with brain MRI, clinical and genetic information, and longitudinal follow-up. We found that hypertension was the most consistent and by far strongest vascular risk factor for WMH components, especially fronto-parietal (C1) and anterior PV (C3) WMH. Higher WHR and BMI were associated with anterior PV (C3) and temporal and deep peri basal ganglia (C6, C7) WMH respectively. A genetic risk score for total WMH volume was associated with most WMH spatial components, most strongly with anterior PV (C3) WMH, but did not show any association with juxtacortical-deep occipital (C5) and deep peri basal ganglia (C7) WMH. Fronto-parietal (C1) and anterior PV (C3) WMH showed the strongest association with general cognitive performance and its decline over time. Anterior PV WMH were also associated with an increased risk of incident all-cause dementia and stroke, with a two-to four-fold risk for participants in the top quartile of the distribution. Larger juxtacortical-deep occipital (C5) WMH volume was predictive of all-cause dementia and intracerebral hemorrhage, but not ischemic stroke. These results suggest that WMH spatial patterns may, at least in part, reflect distinct underlying pathological processes and could, if properly taken into account, prove useful for risk stratification towards personalized treatment of cSVD and prevention of its complications.

Overall, the most striking difference in associations with risk factors and clinical outcomes was observed between, on the one hand, fronto-parietal (C1) and anterior PV (C3) WMH and, on the other hand, juxtacortical-deep occipital (C5) WMH. Anterior PV (C3) WMH and to a lesser extent fronto-parietal WMH (C1) were prominently associated with high blood pressure, genetic risk of total WMH volume, baseline and decline in general cognitive performance, and increased risk of all-cause dementia and both ischemic stroke and intracerebral hemorrhage. In contrast, juxtacortical-deep occipital (C5) WMH showed no evidence of association with vascular or genetic risk factors and was associated with increased risk of intracerebral hemorrhage only. Although pathological evidence would be required for confirmation, this could suggest that fronto-parietal and anterior PV WMH are more likely to reflect arteriosclerotic (and possibly atherosclerotic) processes underlying cSVD, while juxtacortical-deep occipital WMH could be a marker of underlying CAA. Indeed, consistent with this hypothesis, previous studies have reported posterior and juxtacortical WMH distributions in patients with CAA, a subtype of cSVD in which amyloid peptides are deposited in leptomeningeal and cortical vessel walls [64,65]. Of note, posterior WMH have also been associated with parenchymal brain amyloid deposition [15,66] and a study in asymptomatic autosomal dominant AD mutation carriers found occipital and parietal WMH volume to increase two decades before symptom onset [67]. Occipital WMH volume defined using the Bullseye segmentation was higher in amyloid-positive individuals with subjective cognitive decline compared to healthy controls [68]. It has been hypothesized that amyloid peptide might impact pericyte function and damage the neurovascular unit, leading to disrupted blood flow and hypoperfusion [69,70]. Moreover, vascular changes such as stiffness and reduced pulsatile motion of arteries may impair the drainage of amyloid peptides from perivascular space and thereby promote amyloid deposition [71,72]. Hence the association patterns we observed with juxtacortical-deep occipital (C5) WMH might reflect the intricate relationship between vascular and amyloid brain pathology, in contrast with arteriosclerotic cSVD possibly underlying the other components C1 and C3.

The strongest association of high blood pressure with anterior PV (C3) WMH is consistent with previous findings [27,73]. This region (periventricular frontal and peri basal ganglia) are supplied by lenticulostriate arteries which branch from the middle cerebral artery and extend to the periventricular space with very thin arterioles [74]. Compared to vessels supplying the subcortex, ambient blood pressure is high in these small arterioles, which might therefore be more susceptible to lipohyalinosis and arteriosclerotic changes [31,75]. The other vascular risk factors associated with WMH spatial patterns were WHR and BMI with anterior PV and with deep peri basal ganglia and temporal WMH respectively (C3, C6 and C7). Previous studies have reported associations of WHR/BMI with deep WMH primarily, without specifying the lobar location [8,28]. The consistent association of BMI with deep peri basal ganglia WMH in 3C-Dijon and LIFE-Adult might indicate an additional burden of obesity-related systemic inflammation on deep perforating arteries around the basal ganglia. A weighted genetic risk score for total WMH volume was associated with most WMH components except with juxtacortical-deep occipital (C5) and deep basal ganglia (C7) WMH, and most significantly, as for BP traits, with anterior PV WMH. This is consistent with the fact that 60% of genetic risk variants for total WMH volume are also associated with BP [23], and that the anterior PV WMH component had strongest associations with total WMH volume in both cohorts. It also highlights that current understanding of genetic determinants of WMH is likely limited to the most common sub-distributions only, with a knowledge gap for genomics of WMH spatial patterns most likely to reflect underlying CAA [76]. In contrast with previous reports we didn’t find any association of APOE-ε4 or ε2 carrier status with posterior WMH distribution or total WMH volume [8,34], possibly related to insufficient power (significant associations were reported in very large samples of >10,000 participants [23]).

Although different data-driven methods have been applied to study WMH spatial patterns [25–27], no method to date has been exploited to identify molecular mechanisms and enhance disease subtyping, as was done for MRI phenotypes in AD [77]. This is potentially due to difficulties in transferring data driven methods between cohorts which show different distributions of total WMH volume. However, such subtyping would be highly informative to enhance the efficiency of genomics-driven drug discovery through more accurate and cSVD “endotype”-specific phenotype definitions.

Finally, while some associations with clinical outcomes were shared across spatial WMH component, some notable differences were observed. Our preliminary data indeed suggests that juxtacortical-deep occipital WMH might be specifically related to an increased risk of intracerebral hemorrhage, but not ischemic stroke. This suggests that taking into account the spatial distribution of WMH could have important implications for patient stratification towards precision prevention approaches, by approaches, by identifying individuals with covert cSVD at highest risk of developing certain complications and most likely to respond or to develop side-effects from certain investigational therapies. This could for instance be relevant for future trials aiming to explore the efficacy and safety of certain antiplatelet agents in stroke-free persons with covert cSVD. European guidelines currently advise against antiplatelet use in covert cSVD in the absence of other indications [3], due to an increased risk of both ischemic stroke and intracerebral hemorrhage and in the current absence of evidence for benefit in patient subgroups.

We acknowledge limitations. We conducted this study in two population-based cohorts of older individuals with different MRI acquisition parameters. Despite these differences we could however demonstrate the consistency of results regarding vascular and genetic risk, as well as complementary evidence from cognitive analyses. Moreover, we used latent class mixed models to address challenges in longitudinal studies of cognitive function and latent constructs, which allowed us to compare cognitive function even though different tests were used. We did not assess longitudinal change of WMH spatial patterns. This would be interesting to determine how potentially different underlying pathological mechanisms evolve and impact clinical outcomes, especially change in cognitive performance. Moreover, we did not have cerebrospinal fluid or positron-emission tomography based measures of amyloid pathology to indirectly characterize pathological correlates of the different spatial WMH patterns. Finally, both cohorts included participants of mostly European ancestry. To ensure generalizability and enhance impact, future studies should be performed in more diverse samples, including from other ancestry backgrounds [78].

In summary, our findings suggest that using agnostic data-driven approaches in a population-based setting is useful to identify WMH spatial patterns with different risk factor profiles and clinical outcome associations. The findings obtained in two independent cohorts also suggest that the spatial patterns we derived are reproducible. Future studies are needed to decipher the molecular, cellular and pathological underpinnings of WMH spatial components to inform stratification of cSVD patients and pave the way towards precision prevention and therapeutic approaches.

## 8 Acknowledgements

We thank all participants of the 3C-Dijon and LIFE-Adult studies and everyone involved in making these studies possible.

The authors would like to thank Gerard Sanroma for providing the code for deriving the Bullseye segmentation from individual FreeSurfer segmentations openly on https://github.com/gsanroma/bullseye_pipeline.

## 12 Data and Code availability

Due to potential identifiability of individuals from demographic and medical information, we cannot share the data used in this study openly.

The Three-City study is managed by the UMR1219, Bordeaux Population Health Research Center, Bordeaux University, France. Data coming from the Three-City study can be made freely available to interested researchers upon request: http://www.three-citystudy.com/the-three-city-study.php. Contact email address.

Raw data from the LIFE-Adult cohort can be requested via the LIFE data center (https://ldp.life.uni-leipzig.de/).

All code used for analyses in this manuscript is available on https://gitlab.gwdg.de/frauke.beyer/multivariate_risk_svd.

