## Supplemental Material for "White matter hyperintensity spatial patterns: risk factors and clinical correlates"

### Supplementary material for "White matter hyperintensity spatial patterns: risk factors and clinical correlates"

#### 1. Using LIFE-Adult as reference cohort for PCA decomposition

In order to check whether the spatial components were stable across cohorts, we also used LIFE-Adult as reference cohort to derive the spatial components with PCA.

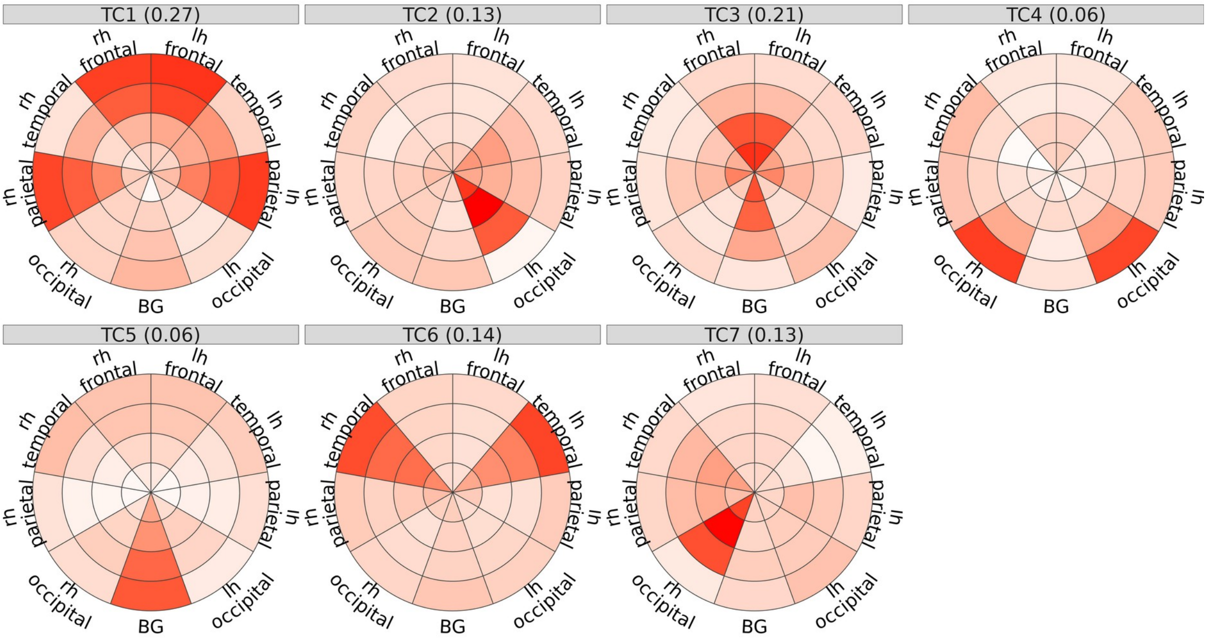


**Figure 1**: WMH components derived from LIFE-Adult using 7 components and oblimin rotation

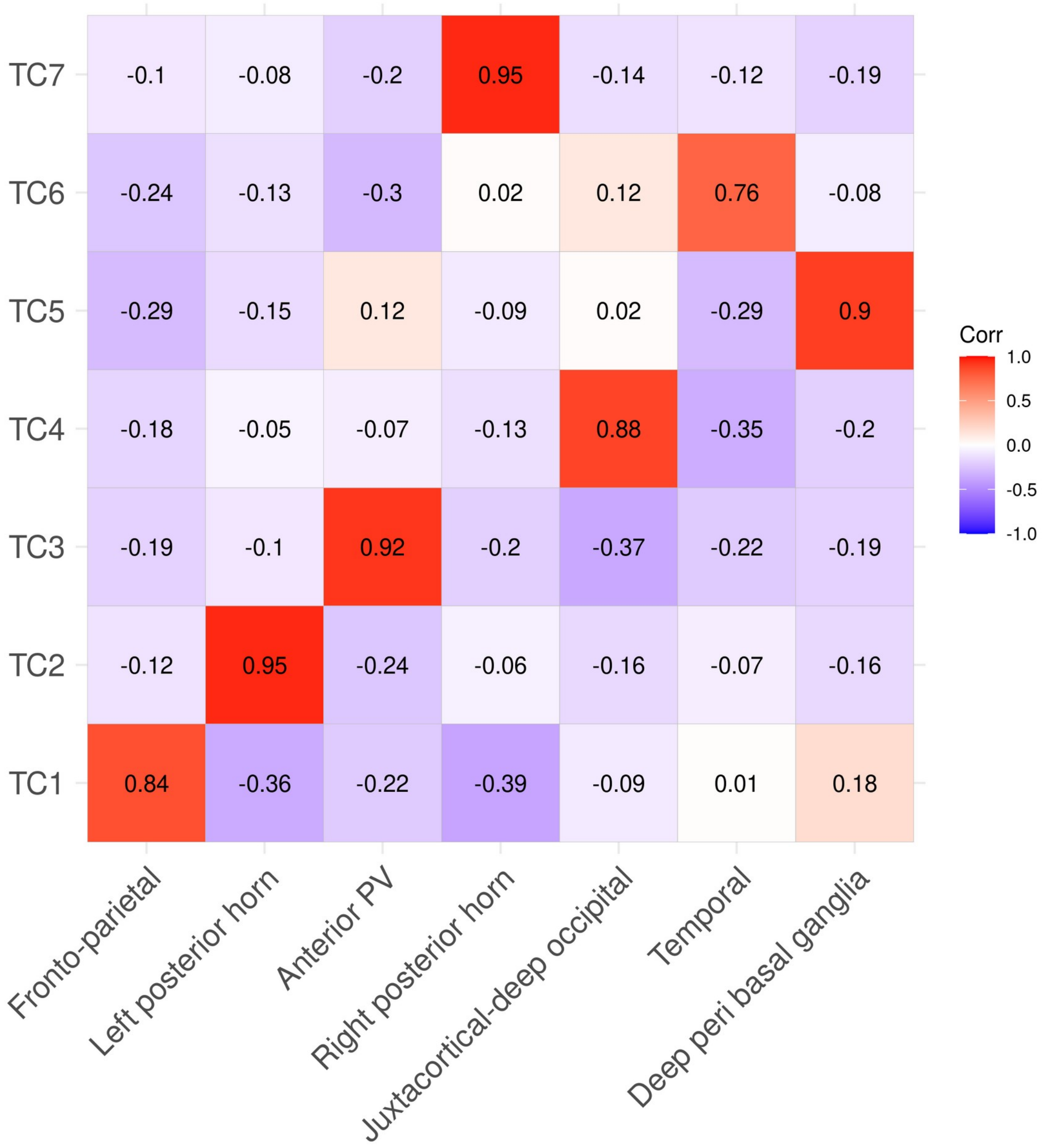


**Figure 2**: Spatial correlation of the components, calculated as correlation of loadings over the 36 Bullseye regions. Anatomical labeling on x-axis refers to components based on 3C-Dijon and described in the manuscript. Numerical labels (TC) on y-axis refer to components based on LIFE-Adult.

For the PCA in LIFE-Adult, the optimal number of components was 5, but we used 7 components to be able to compare to 3C-Dijon. We then calculated the correlation of component loadings between the original components derived from 3C-Dijon (Figure 2, labeled with anatomical descriptions) and the components derived from LIFE-Adult (labeled with numbers, compare Figure 2). The resulting components were very highly correlated (Pearson's $r > 0.75$) with slight changes in the order. We then calculated the correlation of the original and the projected component scores over participants for each sample and component. The results for each component are shown in Figure 3. For all components, both approaches yielded very consistent component scores ($R^2$>0.75). We therefore believe that it was appropriate to use 3C-Dijon as the reference in this study, and that we could expect similar results when deriving components from LIFE-Adult.

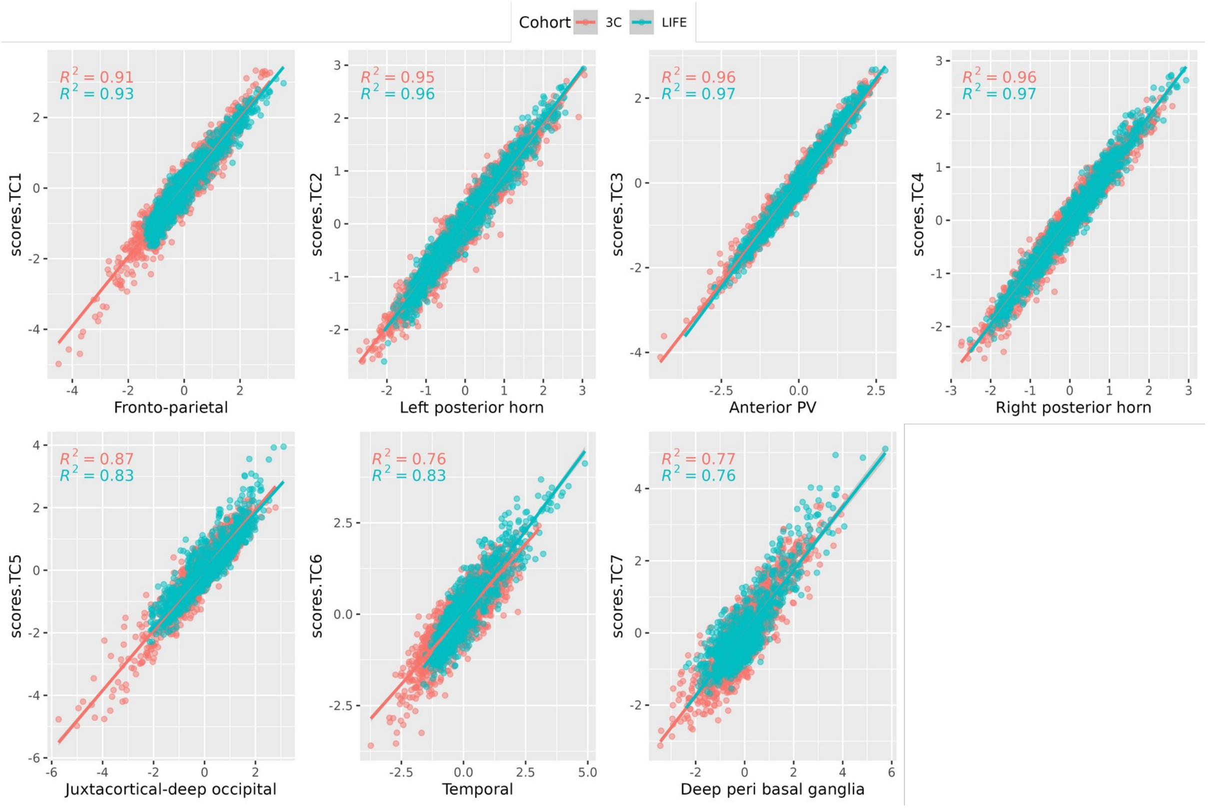


**Figure 3:** Comparison of component scores derived from 3C and LIFE-Adult and their correlations across participants from both cohorts

#### 2 Characterization of WMH spatial patterns

In the following, we aimed to characterize the WMH components derived from 3C-Dijon based on their intercorrelations, and their associations with age and sex.
WMH spatial components were correlated with one another (see Figure 4), as expected when using oblimin rotation in PCA. In general, correlations were weaker in 3C-Dijon. For both cohorts, strongest correlations were between fronto-parietal, anterior PV and temporal WMH. Anterior PV WMH were most strongly associated with total WMH volume (r=0.91 in LIFE-Adult). Left and right posterior horn WMH were strongly correlated in LIFE-Adult and less so in 3C-Dijon. The correlations of juxtacortical-deep occipital and deep basal ganglia WMH with the other components and total WMH were weaker in both studies.

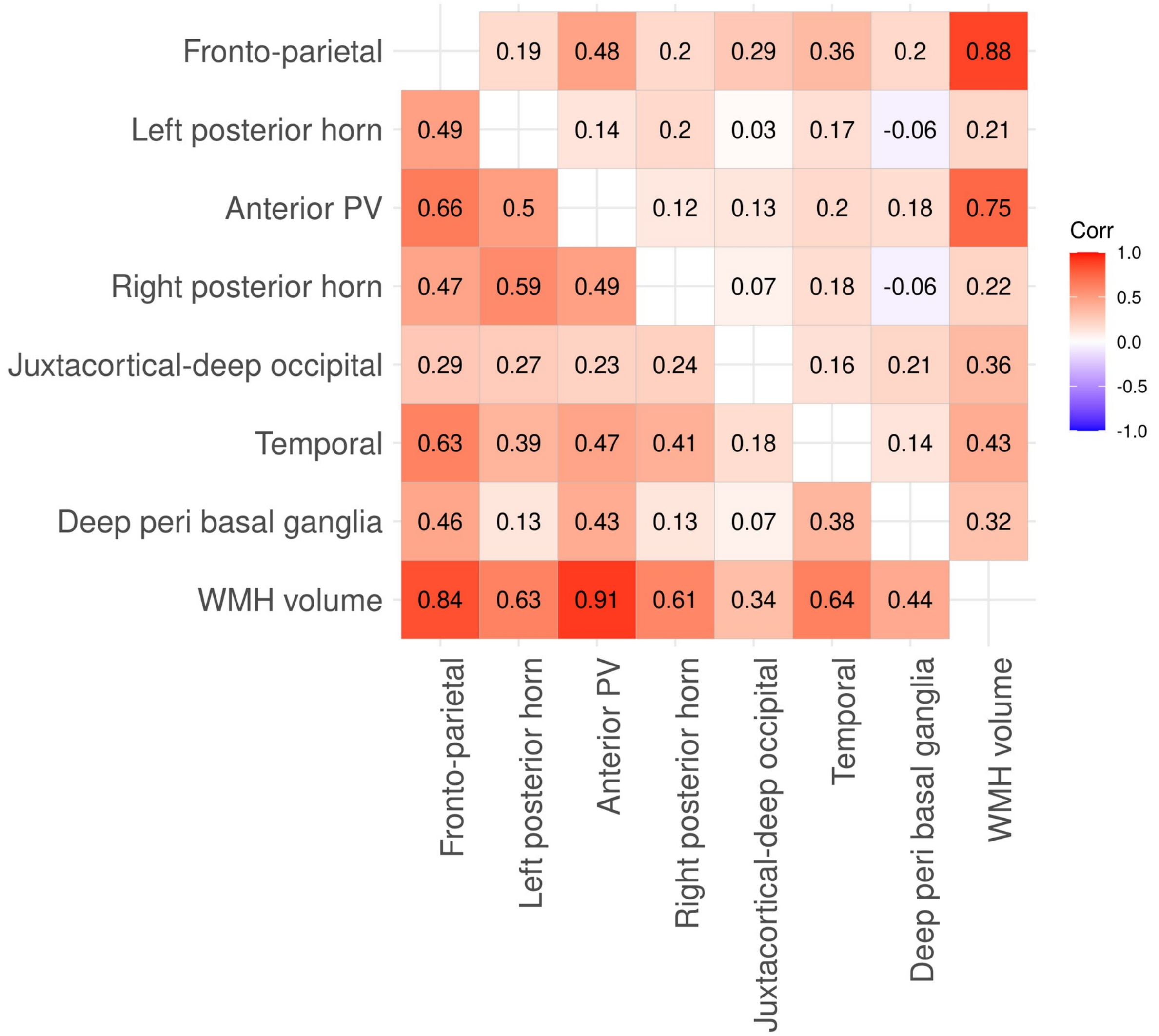

***Figure 4:*** *Correlation of component scores and total WMH volume in 3C-Dijon (upper triangle) and LIFE-Adult (lower triangle)*

### 3 Age- and sex association of WMH spatial components

In 3C-Dijon, only fronto-parietal (C1) and anterior PV WMH (C3) were positively associated with age. All other components except for left posterior horn WMH (C2) showed slight negative associations (Figure 5). In LIFE-Adult, all components were significantly positively associated with age (Figure 6). Age explained least variance in juxtacortical-deep occipital WMH (C5, 1%) and most in anterior PV WMH (C3, 11%). In 3C, males had higher scores in fronto-parietal (C1), right posterior horn (C4) and deep peri basal ganglia WMH (C7) (Figure 7). Similarly, in LIFE, males had higher scores in left and right posterior horn (C2, C4) and temporal WMH (C6) (Figure 8).

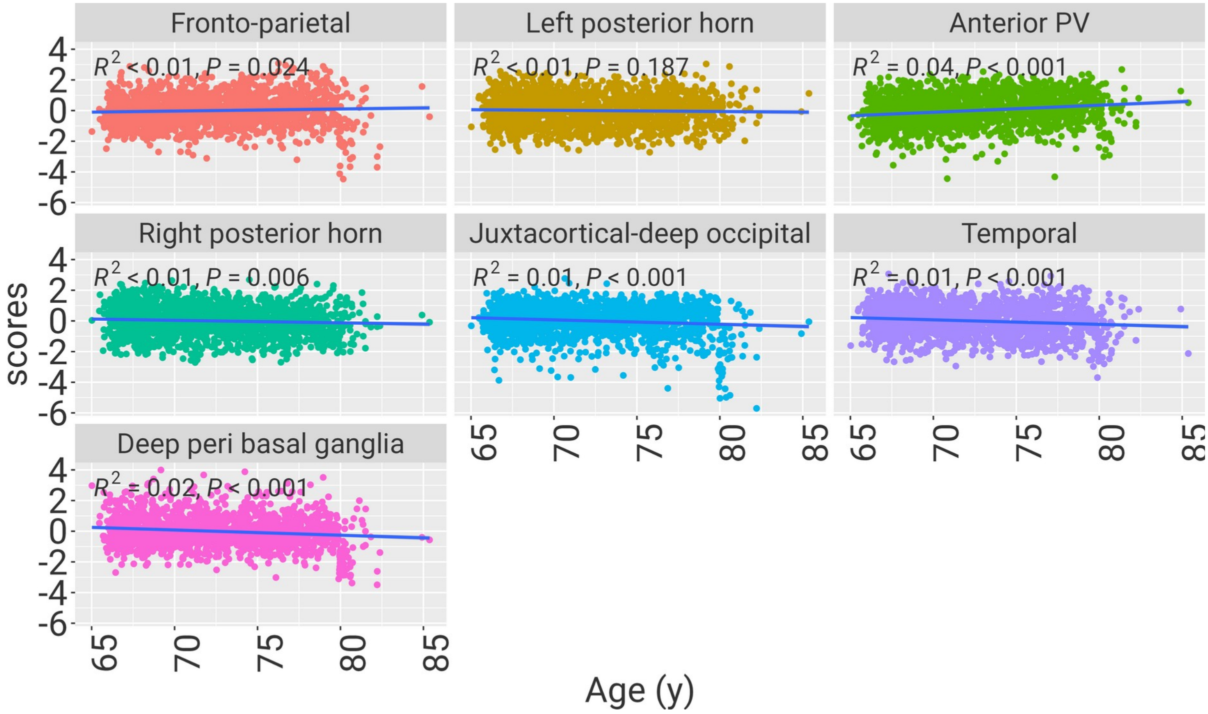


**Figure 5:** Association of age with WMH components in 3C-Dijon. $R^2$ and P-values of simple regression are shown.

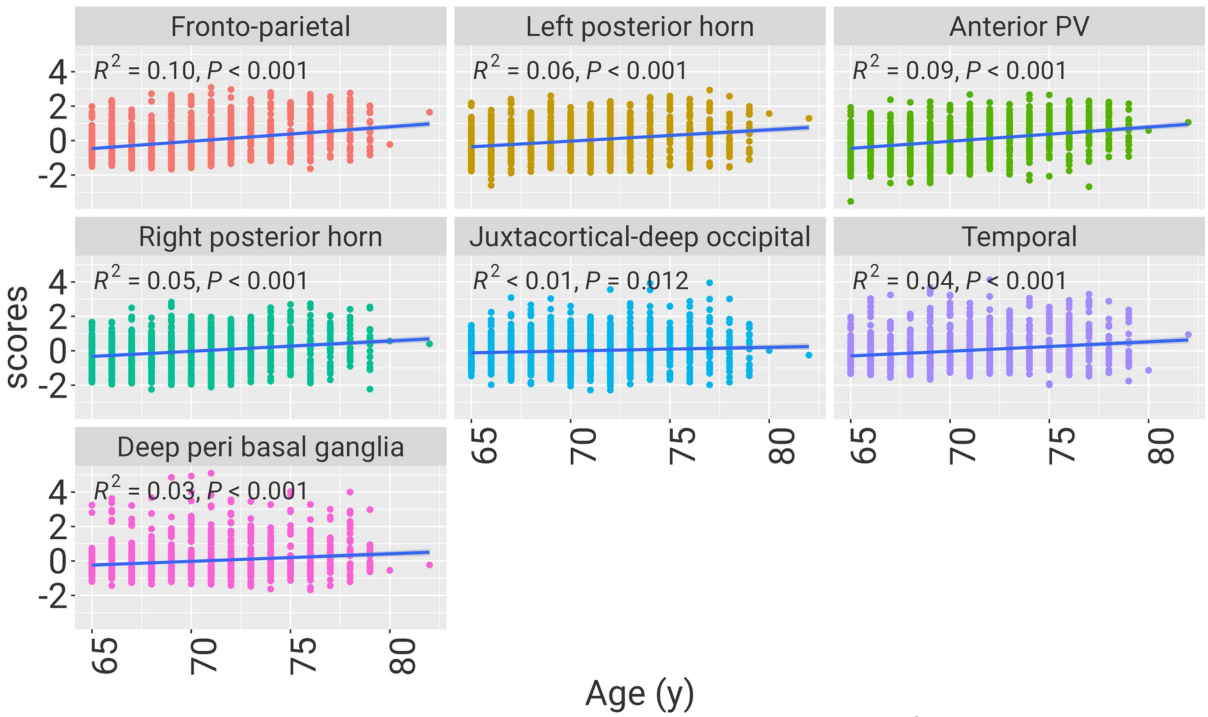


***Figure 6:*** *Association of age with WMH components in LIFE-Adult.* $R^2$ *and P-values of simple regression are shown.*

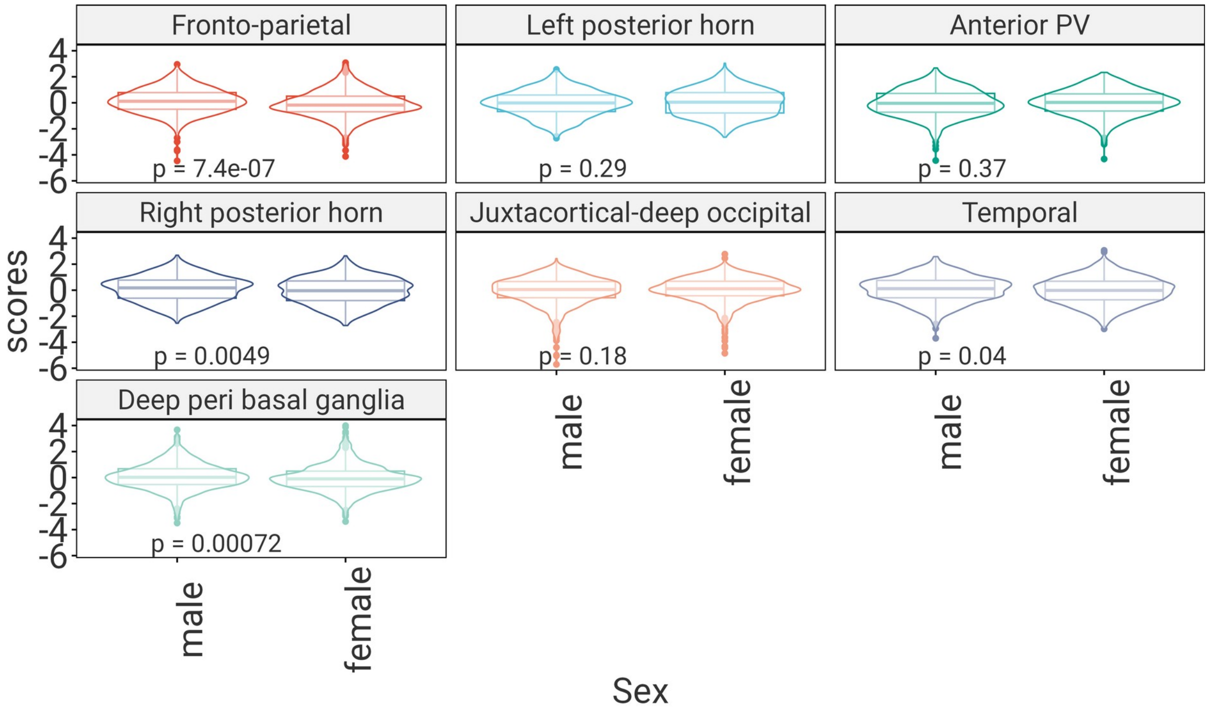


***Figure 7:****Association of sex with WMH components in 3C-Dijon. P-values of two-sample t-tests are shown*

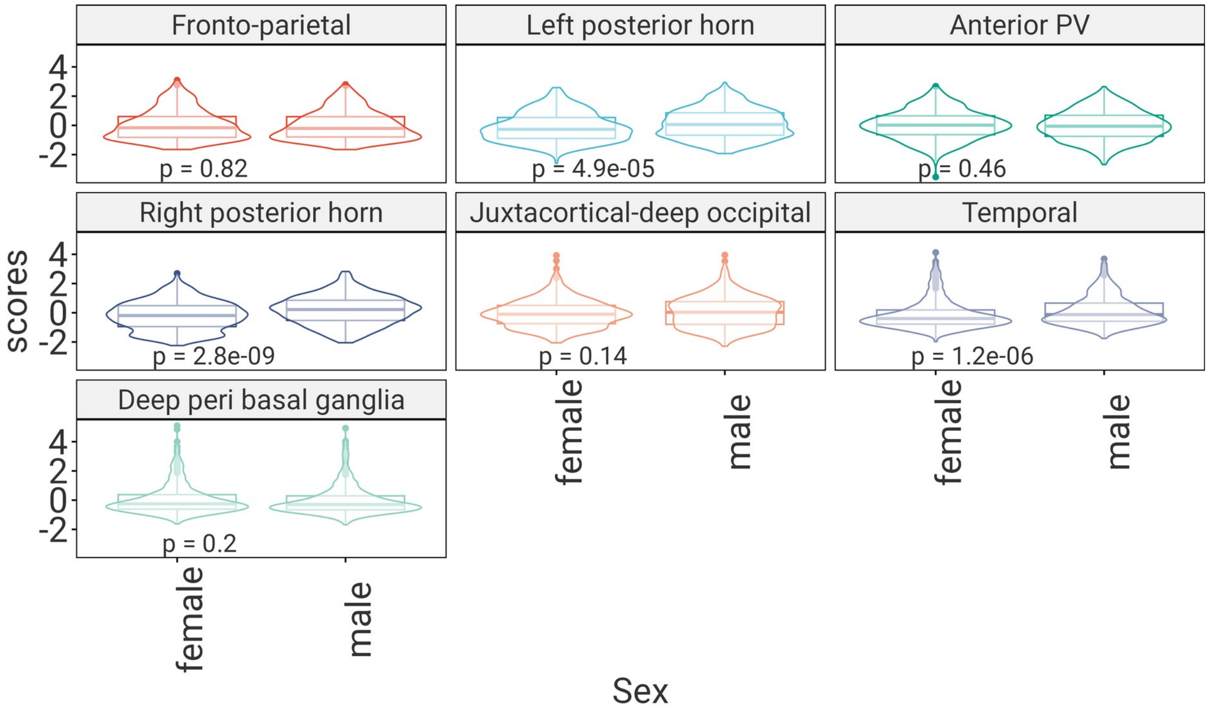


***Figure 8:****Association of sex with WMH components in LIFE-Adult- P-values of two-sample t-tests are shown*

#### 4. Flowcharts

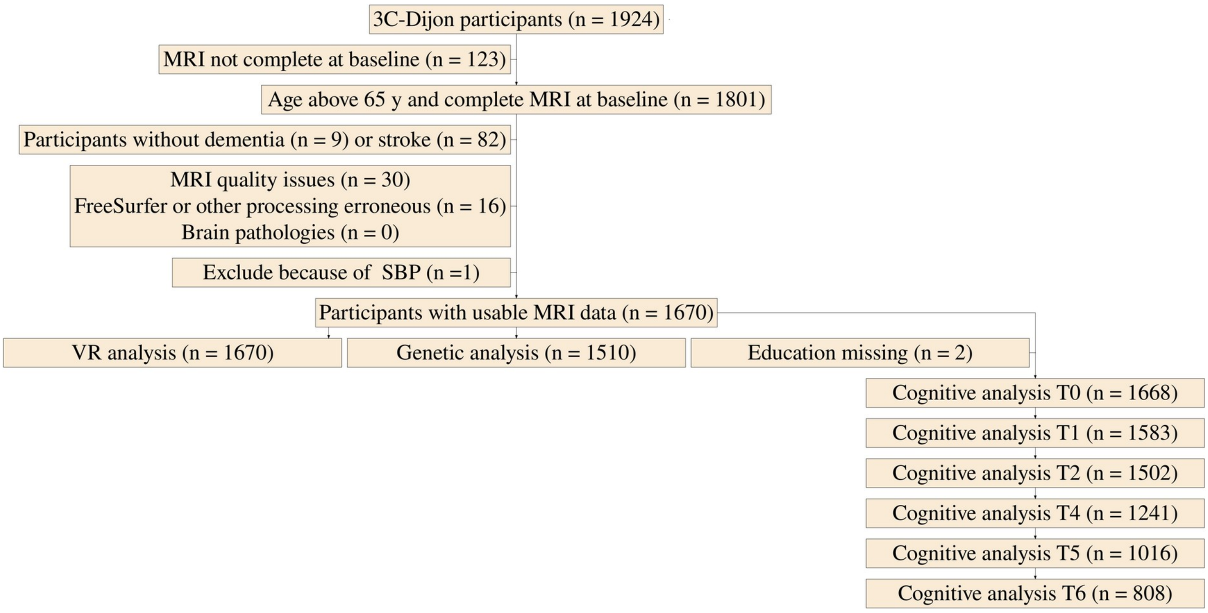


*Figure 9: Flowchart of sample selection for 3C-Dijon*

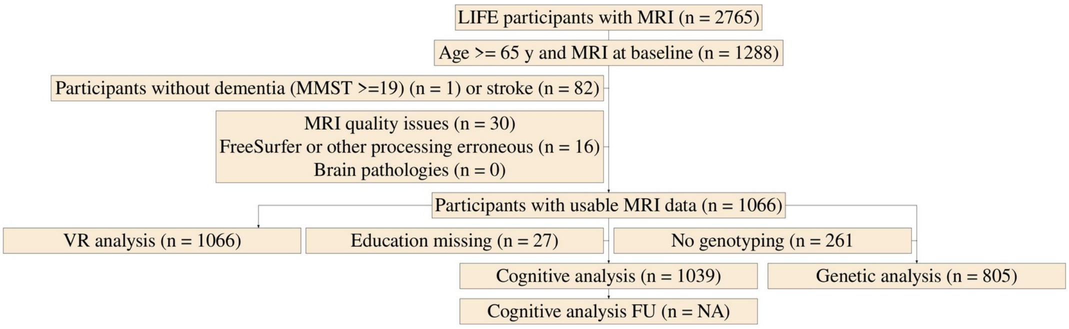


Figure 10: Flowchart of sample selection for LIFE-Adult

#### 5. Details on cognitive assessments in 3C-Dijon and LIFE-Adult

A description of number of participants at baseline and throughout waves in 3C-Dijon is found in Tables 1 and 2. The sample for the baseline cognitive assessment in LIFE-Adult is described in Table 3.

*Table 1: Characteristics of baseline cognitive sample in 3C-Dijon*

| Variable | N | Mean/% | Std. Dev. |
|---|---|---|---|
| Age (y) | 1668 | 72 | 4.1 |
| Gender(women) | 1668 (1011) | 61% | |
| Education | | | |
| no formal education | 85 | 5% | |
| primary education with diploma | 179 | 11% | |
| secondary education short | 733 | 44% | |
| secondary education long | 313 | 19% | |
| higher education | 358 | 21% | |

*Table 2: Characteristics of cognitive assessment across waves in 3C-Dijon. Shown are average, standard deviation, minimal and maximal values of raw test scores as well as average time since baseline per time point. N indicates number of participants who participated in each of the assessments at timepoints T0, T1, T2, T4, T5 and T6.*

| Variable | N | Mean | SD | Min | Max |
|---|---|---|---|---|---|
| **T0 (Time since baseline in years)** | 1668 | 0 | 0 | 0 | 0 |
| BENTON | 1655 | 11.8 | 1.88 | 0 | 15 |
| ISA | 1659 | 34.2 | 6.73 | 11 | 60 |
| TMTA | 1651 | 52.6 | 20.1 | 15 | 300 |
| TMTB | 1621 | 101 | 41.1 | 11 | 300 |
| **T1** | 1583 | 1.85 | 0.172 | 0.751 | 3.66 |
| BENTON | 1577 | 11.7 | 2.02 | 0 | 15 |
| ISA | 1583 | 36.4 | 6.78 | 12 | 59 |
| TMTA | 0 | | | | |
| TMTB | 0 | | | | |
| **T2** | 1502 | 3.58 | 0.206 | 2.83 | 5.4 |
| BENTON | 1495 | 11.4 | 1.99 | 1 | 15 |
| ISA | 1500 | 35.5 | 6.66 | 4 | 66 |
| TMTA | 1482 | 53.1 | 19.9 | 18 | 210 |
| TMTB | 1462 | 104 | 43.7 | 18 | 300 |
| **T4** | 1241 | 7.04 | 0.379 | 5.35 | 8.61 |
| BENTON | 1237 | 11.6 | 2.21 | 0 | 15 |
| ISA | 1240 | 34.6 | 6.78 | 9 | 58 |
| TMTA | 1228 | 48.6 | 18.8 | 16 | 200 |
| TMTB | 1219 | 119 | 48.3 | 16 | 300 |
| **T5** | 1016 | 8.8 | 0.413 | 7.35 | 10.6 |
| BENTON | 1013 | 11.9 | 2.09 | 0 | 15 |
| ISA | 1016 | 35.2 | 6.88 | 13 | 64 |
| TMTA | 1006 | 48.6 | 20.3 | 20 | 206 |
| TMTB | 997 | 118 | 47.8 | 22 | 300 |

| Variable | N | Mean | SD | Min | Max |
|---|---|---|---|---|---|
| **T6** | 808 | 10.8 | 0.387 | 9.67 | 12.2 |
| BENTON | 796 | 12.1 | 1.9 | 0 | 15 |
| ISA | 805 | 35 | 6.46 | 13 | 55 |
| TMTA | 801 | 48.4 | 17.6 | 16 | 154 |
| TMTB | 792 | 117 | 47.5 | 43 | 300 |

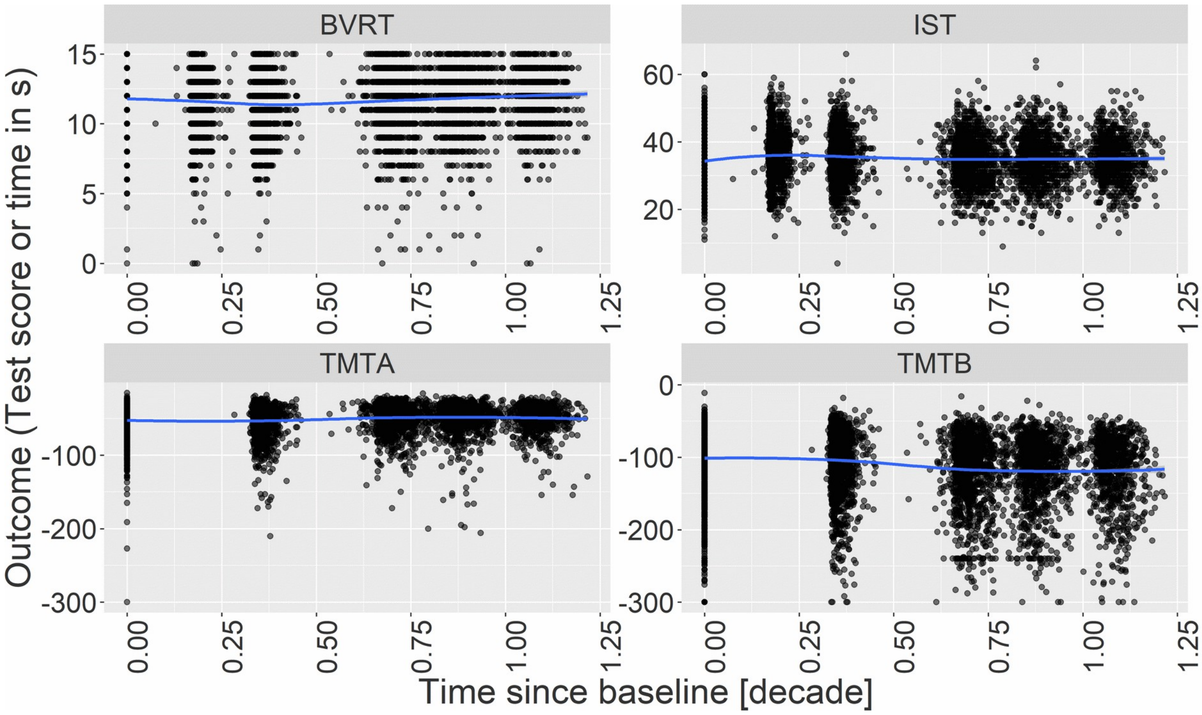


*Figure 11: Overview of trajectories of neuropsychological test scores in 3C-Dijon. Blue line shows local polynomial regression.*

*Table 3: Descriptive summary of the subsample with cognitive data in LIFE-Adult*

| Variable | N | Mean/% | Median |
|---|---|---|---|
| Age (y) | 1039 | 70.5 | 70 |
| Gender (women) | 1039 (475) | 45.7% | |
| Education | | | |
| no secondary school degree, | 7 | 0.7% | |
| secondary school degree, | 86 | 8.3% | |
| advanced secondary school degree | 576 | 55.4% | |
| university entrance degree | 370 | 35.6% | |
| MMST | 1020 | 28.5 | 29 |

#### 5. Vascular risk factor results

*Table 4: Associations of VR factors with WMH spatial components in the 3C-Dijon and LIFE-Adult samples. Shown are all associations with pFDR<0.05 in at least one of the cohorts. Estimate refers to standardized regression estimates from regression models adjusting for age, sex and TIV. FDR-p values are across cohorts and estimates. β, SE, and p-value from the meta-analysis. APOE-e2: dominant model, APOE-e2_homo: comparing homozygotes to non-carriers, APOE-e2_33: comparing homozygotes to 33 carriers, HTN: hypertension, SBP: systolic blood pressure, DBP: diastolic blood pressure, WHR: waist-to-hip ratio, WMH: white matter hyperintensities, FDR: false-discovery rate, SE: standard error.*

| | | 3C-Dijon | | | LIFE | | | Meta-analysis | | |
|---|---|---|---|---|---|---|---|---|---|---|
| **WMH Measure** | **Predictor** | **β** | **SE** | **pFDR** | **β** | **SE** | **pFDR** | **β** | **SE** | **p** |
| Fronto-Parietal | HTN | 0.173 | 0.058 | 0.7 | 0.251 | 0.062 | 8.08e-04 | 0.210 | 0.042 | 8.18e-07 |
| Fronto-Parietal | SBP | 0.022 | 0.025 | 0.2 | 0.171 | 0.030 | 6.98e-07 | 0.084 | 0.019 | 1.11e-05 |
| Fronto-Parietal | DBP | 0.096 | 0.024 | 0.26 | 0.157 | 0.030 | 6.24e-06 | 0.121 | 0.019 | 1.84e-10 |
| Fronto-Parietal | BMI | 0.014 | 0.024 | 0.035 | 0.073 | 0.029 | 0.073 | 0.038 | 0.019 | 0.041 |
| Fronto-Parietal | WHR | 0.007 | 0.024 | 0.91 | 0.085 | 0.029 | 0.031 | 0.039 | 0.019 | 0.037 |
| Fronto-Parietal | WHR binary | 0.036 | 0.056 | 0.011 | 0.183 | 0.088 | 0.15 | 0.079 | 0.047 | 0.096 |
| Fronto-Parietal | Diabetes | 0.068 | 0.077 | 0.011 | 0.131 | 0.073 | 0.23 | 0.101 | 0.053 | 0.057 |
| Left Posterior | SBP | -0.003 | 0.026 | 0.2 | 0.125 | 0.030 | 6.25e-04 | 0.051 | 0.019 | 0.0095 |

| | | 3C-Dijon | | | LIFE | | | Meta-analysis | | |
|---|---|---|---|---|---|---|---|---|---|---|
| **WMH Measure** | **Predictor** | **β** | **SE** | **pFDR** | **β** | **SE** | **pFDR** | **β** | **SE** | **p** |
| Horn | | | | | | | | | | |
| Left Posterior Horn | DBP | 0.020 | 0.025 | 0.43 | 0.103 | 0.030 | 0.008 | 0.054 | 0.019 | 0.0055 |
| Left Posterior Horn | Smoking | -0.046 | 0.062 | 0.86 | 0.011 | 0.069 | 0.0077 | -0.021 | 0.046 | 0.65 |
| Anterior PV | HTN | 0.238 | 0.058 | 0.95 | 0.298 | 0.062 | 4.49e-05 | 0.266 | 0.042 | 3.33e-10 |
| Anterior PV | DBP | 0.149 | 0.024 | 0.74 | 0.154 | 0.030 | 1.13e-05 | 0.151 | 0.019 | 1.33e-15 |
| Anterior PV | WHR | 0.061 | 0.024 | 0.82 | 0.094 | 0.029 | 0.011 | 0.075 | 0.019 | 6.06e-05 |
| Anterior PV | Smoking | 0.057 | 0.061 | 0.76 | 0.067 | 0.069 | 7.20e-04 | 0.062 | 0.046 | 0.18 |
| Right posterior horn | SBP | -0.038 | 0.025 | 0.41 | 0.110 | 0.030 | 0.0032 | 0.023 | 0.019 | 0.23 |
| Right posterior horn | DBP | -0.002 | 0.025 | 0.57 | 0.116 | 0.030 | 0.0018 | 0.046 | 0.019 | 0.018 |
| Right posterior horn | Smoking | 0.046 | 0.062 | 0.38 | -0.099 | 0.069 | 4.49e-05 | -0.019 | 0.046 | 0.69 |
| Juxtacortical-deep occipital | Diabetes | 0.135 | 0.078 | 0.7 | -0.059 | 0.077 | 8.08e-04 | 0.037 | 0.055 | 0.5 |
| Juxtacortical-deep occipital | Smoking | 0.078 | 0.062 | 0.2 | -0.033 | 0.072 | 6.98e-07 | 0.031 | 0.047 | 0.51 |

| | | 3C-Dijon | | | LIFE | | | Meta-analysis | | |
|---|---|---|---|---|---|---|---|---|---|---|
| **WMH Measure** | **Predictor** | **β** | **SE** | **pFDR** | **β** | **SE** | **pFDR** | **β** | **SE** | **p** |
| Juxtacortical-deep occipital | APOE-e2 | 0.056 | 0.070 | 0.035 | -0.263 | 0.092 | 0.073 | -0.061 | 0.056 | 0.28 |
| Juxtacortical-deep occipital | APOE-e2_homo | 0.201 | 0.335 | 0.011 | -1.163 | 0.358 | 0.15 | -0.436 | 0.245 | 0.075 |
| Juxtacortical-deep occipital | APOE-e2_33 | 0.208 | 0.334 | 0.011 | -1.142 | 0.353 | 0.23 | -0.429 | 0.242 | 0.076 |
| Temporal | HTN | 0.120 | 0.059 | 0.86 | 0.214 | 0.063 | 0.0077 | 0.164 | 0.043 | 1.42e-04 |
| Temporal | BMI | 0.045 | 0.025 | 0.9 | 0.110 | 0.029 | 0.0021 | 0.072 | 0.019 | 1.37e-04 |
| Temporal | Smoking | -0.057 | 0.062 | 0.2 | 0.051 | 0.069 | 6.25e-04 | -0.008 | 0.046 | 0.86 |
| Deep peri basal ganglia | HTN | 0.103 | 0.059 | 0.76 | 0.263 | 0.064 | 7.20e-04 | 0.176 | 0.043 | 5.35e-05 |
| Deep peri basal ganglia | Diabetes | 0.170 | 0.078 | 0.95 | 0.007 | 0.076 | 4.49e-05 | 0.087 | 0.054 | 0.11 |
| Deep peri basal ganglia | Smoking | 0.064 | 0.061 | 0.82 | 0.004 | 0.070 | 3.54e-07 | 0.038 | 0.046 | 0.41 |
| WMH volume | HTN | 0.241 | 0.057 | 0.38 | 0.292 | 0.061 | 4.49e-05 | 0.265 | 0.042 | 1.96e-10 |
| WMH volume | SBP | 0.071 | 0.025 | 0.87 | 0.183 | 0.029 | 1.21e-07 | 0.117 | 0.019 | 4.89e-10 |
| WMH volume | DBP | 0.133 | 0.024 | 0.51 | 0.161 | 0.030 | 2.34e-06 | 0.144 | 0.019 | 9.48e-15 |
| WMH volume | WHR | 0.035 | 0.024 | 0.7 | 0.094 | 0.029 | 0.01 | 0.059 | 0.018 | 0.0012 |
| WMH volume | Smoking | 0.048 | 0.060 | 0.41 | 0.009 | 0.067 | 0.0032 | 0.031 | 0.045 | 0.49 |

#### 6. APOE results

*Table 5: Associations of APOE genotypes (APOE-e4/APOE-e2: dominant model including e24 alleles and APOE-e4_homo/APOE-e2_homo: model comparing homozygotes to non-carriers) with WMH spatial components in the 3C and LIFE cohorts. Shown are regression estimates from regression models adjusting for age, sex and TIV, FDR-corrected p-values for both cohorts and estimate, p-value and FDR-p from the meta-analysis. APOE: apoliproprotein E, WMH: white matter hyperintensities, FDR: false-discovery rate, SE: standard error.*

| | | 3C | | | | LIFE | | | | Meta-analysis | | | |
|---|---|---|---|---|---|---|---|---|---|---|---|---|---|
| **WMH Measure** | **Model** | **β** | **SE** | **p** | **pFDR** | **β** | **SE** | **p** | **pFDR** | **β** | **SE** | **p** | **pFDR** |
| Fronto-Parietal | APOE-e4 | -0.03 | 0.06 | 0.65 | 0.81 | -0.02 | 0.07 | 0.79 | 0.89 | -0.02 | 0.04 | 0.61 | 0.78 |
| Left Posterior Horn | APOE-e4 | -0.05 | 0.06 | 0.44 | 0.73 | -0.06 | 0.07 | 0.40 | 0.61 | -0.05 | 0.05 | 0.25 | 0.48 |
| Anterior PV | APOE-e4 | -0.05 | 0.06 | 0.35 | 0.63 | -0.10 | 0.07 | 0.15 | 0.31 | -0.07 | 0.04 | 0.10 | 0.25 |
| Right posterior horn | APOE-e4 | 0.02 | 0.06 | 0.73 | 0.85 | 0.02 | 0.07 | 0.76 | 0.88 | 0.02 | 0.05 | 0.65 | 0.80 |
| Juxtacortical-deep occipital | APOE-e4 | -0.03 | 0.06 | 0.63 | 0.81 | 0.12 | 0.07 | 0.10 | 0.22 | 0.03 | 0.05 | 0.52 | 0.70 |
| Temporal | APOE-e4 | 0.03 | 0.06 | 0.66 | 0.81 | 0.09 | 0.07 | 0.23 | 0.40 | 0.05 | 0.05 | 0.27 | 0.48 |
| Deep peri basal ganglia | APOE-e4 | -0.13 | 0.06 | 0.02 | 0.14 | -0.05 | 0.07 | 0.52 | 0.73 | -0.10 | 0.05 | 0.03 | 0.09 |
| WMH volume | APOE-e4 | -0.04 | 0.06 | 0.51 | 0.76 | -0.07 | 0.07 | 0.33 | 0.52 | -0.05 | 0.04 | 0.26 | 0.48 |
| Fronto-Parietal | APOE-e2 | 0.14 | 0.07 | 0.04 | 0.23 | -0.20 | 0.09 | 0.02 | 0.08 | 0.01 | 0.05 | 0.88 | 0.92 |
| Left Posterior Horn | APOE-e2 | -0.03 | 0.07 | 0.71 | 0.84 | -0.12 | 0.09 | 0.19 | 0.36 | -0.06 | 0.06 | 0.27 | 0.48 |
| Anterior PV | APOE-e2 | 0.08 | 0.07 | 0.25 | 0.49 | -0.12 | 0.09 | 0.19 | 0.36 | 0.01 | 0.05 | 0.91 | 0.94 |
| Right posterior horn | APOE-e2 | 0.05 | 0.07 | 0.52 | 0.76 | -0.09 | 0.09 | 0.29 | 0.48 | -0.01 | 0.06 | 0.88 | 0.92 |
| Juxtacortical-deep occipital | APOE-e2 | 0.06 | 0.07 | 0.43 | 0.72 | -0.26 | 0.09 | 0.00 | 0.02 | -0.06 | 0.06 | 0.28 | 0.48 |
| Temporal | APOE-e2 | 0.09 | 0.07 | 0.18 | 0.39 | -0.02 | 0.09 | 0.87 | 0.90 | 0.05 | 0.06 | 0.34 | 0.56 |
| Deep peri basal ganglia | APOE-e2 | 0.04 | 0.07 | 0.59 | 0.78 | -0.02 | 0.09 | 0.85 | 0.90 | 0.02 | 0.06 | 0.76 | 0.87 |
| WMH volume | APOE-e2 | 0.13 | 0.07 | 0.06 | 0.26 | -0.15 | 0.09 | 0.09 | 0.21 | 0.02 | 0.05 | 0.65 | 0.80 |
| Fronto-Parietal | APOE-e4_homo | 0.49 | 0.21 | 0.02 | 0.14 | 0.27 | 0.24 | 0.26 | 0.45 | 0.40 | 0.16 | 0.01 | 0.05 |
| Left Posterior Horn | APOE-e4_homo | 0.13 | 0.22 | 0.54 | 0.76 | -0.05 | 0.24 | 0.83 | 0.89 | 0.05 | 0.16 | 0.76 | 0.87 |
| Anterior PV | APOE-e4_homo | 0.09 | 0.21 | 0.68 | 0.82 | -0.13 | 0.24 | 0.59 | 0.78 | -0.01 | 0.16 | 0.96 | 0.97 |

| | | 3C | | | | LIFE | | | | Meta-analysis | | | |
|---|---|---|---|---|---|---|---|---|---|---|---|---|---|
| **WMH Measure** | **Model** | **β** | **SE** | **p** | **pFDR** | **β** | **SE** | **p** | **pFDR** | **β** | **SE** | **p** | **pFDR** |
| Right posterior horn | APOE-e4_homo | 0.08 | 0.22 | 0.72 | 0.85 | 0.47 | 0.24 | 0.05 | 0.15 | 0.25 | 0.16 | 0.12 | 0.27 |
| Juxtacortical-deep occipital | APOE-e4_homo | 0.33 | 0.22 | 0.13 | 0.35 | 0.05 | 0.25 | 0.84 | 0.90 | 0.21 | 0.16 | 0.20 | 0.41 |
| Temporal | APOE-e4_homo | 0.31 | 0.22 | 0.16 | 0.37 | 0.34 | 0.25 | 0.17 | 0.33 | 0.32 | 0.16 | 0.05 | 0.13 |
| Deep peri basal ganglia | APOE-e4_homo | -0.09 | 0.22 | 0.69 | 0.82 | -0.11 | 0.26 | 0.68 | 0.83 | -0.10 | 0.16 | 0.56 | 0.75 |
| WMH volume | APOE-e4_homo | 0.49 | 0.21 | 0.02 | 0.14 | 0.11 | 0.24 | 0.65 | 0.81 | 0.32 | 0.16 | 0.04 | 0.12 |
| Fronto-Parietal | APOE-e2_homo | 0.23 | 0.33 | 0.49 | 0.76 | -0.26 | 0.34 | 0.44 | 0.66 | -0.01 | 0.24 | 0.97 | 0.97 |
| Left Posterior Horn | APOE-e2_homo | -0.49 | 0.33 | 0.14 | 0.36 | 0.05 | 0.34 | 0.88 | 0.91 | -0.22 | 0.24 | 0.35 | 0.56 |
| Anterior PV | APOE-e2_homo | -0.08 | 0.33 | 0.80 | 0.88 | -0.35 | 0.35 | 0.32 | 0.52 | -0.21 | 0.24 | 0.38 | 0.58 |
| Right posterior horn | APOE-e2_homo | -0.01 | 0.33 | 0.97 | 0.98 | -0.26 | 0.34 | 0.45 | 0.66 | -0.13 | 0.24 | 0.58 | 0.77 |
| Juxtacortical-deep occipital | APOE-e2_homo | 0.20 | 0.34 | 0.55 | 0.76 | -1.16 | 0.36 | 0.00 | 0.01 | -0.44 | 0.24 | 0.07 | 0.19 |
| Temporal | APOE-e2_homo | 0.17 | 0.33 | 0.61 | 0.80 | -0.11 | 0.35 | 0.74 | 0.87 | 0.03 | 0.24 | 0.88 | 0.92 |
| Deep peri basal ganglia | APOE-e2_homo | 0.06 | 0.33 | 0.85 | 0.91 | -0.17 | 0.36 | 0.64 | 0.81 | -0.04 | 0.25 | 0.86 | 0.92 |
| WMH volume | APOE-e2_homo | 0.20 | 0.32 | 0.53 | 0.76 | -0.38 | 0.34 | 0.26 | 0.45 | -0.07 | 0.23 | 0.75 | 0.87 |

### 7 Executive function results

*Table 6: Associations of WMH spatial components with baseline and change in executive function in 3C-Dijon and LIFE. Regression estimates are from the latent class mixed-effect models with WMH components and total WMH volume as predictors for intercept and change of general cognitive function, adjusting for age, sex and education. FDR-corrected p-values were calculated separately per cohort. Also shown are estimates and p-values from the meta-analysis. WMH: white matter hyperintensities, FDR: false-discovery rate, SE: standard error.*

| | | 3C | | | | LIFE | | | | Meta-analysis | | | |
|---|---|---|---|---|---|---|---|---|---|---|---|---|---|
| **WMH Measure** | **Model** | **β** | **SE** | **p** | **pFDR** | **β** | **SE** | **p** | **pFDR** | **β** | **SE** | **p** | **pFDR** |
| Fronto-Parietal | Intercept | -0.105 | 0.033 | 0.00176 | 0.111 | -0.163 | 0.045 | 3.00e-04 | 0.0255 | -0.125 | 0.027 | 3.02e-06 | 2.42e-05 |
| Fronto-Parietal | Change | -0.012 | 0.044 | 0.789 | 1.000 | -0.070 | 0.080 | 0.38 | 1 | -0.026 | 0.039 | 0.509 | 0.627 |
| Left Posterior Horn | Intercept | 0.018 | 0.032 | 0.583 | 1.000 | -0.090 | 0.044 | 0.0414 | 1 | -0.020 | 0.026 | 0.442 | 0.627 |
| Left Posterior Horn | Change | 0.004 | 0.044 | 0.931 | 1.000 | -0.164 | 0.076 | 0.03 | 1 | -0.039 | 0.038 | 0.31 | 0.495 |
| Anterior PV | Intercept | -0.100 | 0.034 | 0.00295 | 0.180 | -0.121 | 0.045 | 0.00675 | 0.479 | -0.108 | 0.027 | 6.19e-05 | 3.30e-04 |
| Anterior PV | Change | -0.119 | 0.042 | 0.00506 | 0.301 | -0.076 | 0.078 | 0.33 | 1 | -0.109 | 0.037 | 0.00342 | 0.0109 |
| Right posterior horn | Intercept | 0.032 | 0.033 | 0.332 | 1.000 | -0.050 | 0.044 | 0.251 | 1 | 0.002 | 0.026 | 0.933 | 0.933 |

| | | 3C | | | | LIFE | | | | Meta-analysis | | | |
|---|---|---|---|---|---|---|---|---|---|---|---|---|---|
| **WMH Measure** | **Model** | **β** | **SE** | **p** | **pFDR** | **β** | **SE** | **p** | **pFDR** | **β** | **SE** | **p** | **pFDR** |
| Right posterior horn | Change | 0.013 | 0.041 | 0.751 | 1.000 | -0.149 | 0.075 | 0.0482 | 1 | -0.025 | 0.036 | 0.499 | 0.627 |
| Juxtacortical-deep occipital | Intercept | 0.039 | 0.033 | 0.239 | 1.000 | -0.070 | 0.042 | 0.0963 | 1 | -0.003 | 0.026 | 0.917 | 0.933 |
| Juxtacortical-deep occipital | Change | 0.005 | 0.043 | 0.901 | 1.000 | -0.090 | 0.077 | 0.24 | 1 | -0.018 | 0.038 | 0.64 | 0.731 |
| Temporal | Intercept | -0.017 | 0.033 | 0.608 | 1.000 | -0.179 | 0.045 | 6.00e-05 | 0.00522 | -0.075 | 0.027 | 0.00521 | 0.0139 |
| Temporal | Change | 0.059 | 0.043 | 0.169 | 1.000 | 0.013 | 0.081 | 0.874 | 1 | 0.049 | 0.038 | 0.197 | 0.351 |
| Deep peri basal ganglia | Intercept | -0.067 | 0.033 | 0.0422 | 1.000 | -0.149 | 0.043 | 5.40e-04 | 0.0443 | -0.098 | 0.026 | 2.01e-04 | 8.04e-04 |
| Deep peri basal ganglia | Change | 0.080 | 0.042 | 0.0575 | 1.000 | 0.063 | 0.085 | 0.455 | 1 | 0.077 | 0.038 | 0.0419 | 0.0959 |
| WMH volume | Intercept | -0.124 | 0.034 | 2.60e-04 | 0.017 | -0.154 | 0.045 | 7.30e-04 | 0.0584 | -0.135 | 0.027 | 7.41e-07 | 1.19e-05 |
| WMH volume | Change | -0.051 | 0.044 | 0.243 | 1.000 | -0.120 | 0.079 | 0.129 | 1 | -0.068 | 0.038 | 0.0788 | 0.158 |

### 8 Memory function

*Table 7: Associations of WMH spatial components with baseline and change in memory function in 3C-Dijon. Regression estimates are from the latent class mixed-effect models with WMH components and total WMH volume as predictors for intercept and (linear and quadratic) change of visual memory function, adjusting for age, sex and education. WMH: white matter hyperintensities, FDR: false-discovery rate, SE: standard error.*

| WMH Measure | Effect | β | SE | p | pFDR |
|---|---|---|---|---|---|
| Fronto-Parietal | Intercept | -0.032 | 0.028 | 0.259 | 1.000 |
| Fronto-Parietal | Linear | 0.057 | 0.118 | 0.629 | 1.000 |
| Fronto-Parietal | Quadratic | -0.708 | 1.122 | 0.528 | 1.000 |
| Left posterior horn | Intercept | 0.024 | 0.034 | 0.477 | 1.000 |
| Left posterior horn | Linear | -0.005 | 0.196 | 0.981 | 1.000 |
| Left posterior horn | Quadratic | 0.182 | 1.826 | 0.921 | 1.000 |
| Anterior PV | Intercept | -0.054 | 0.028 | 0.054 | 1.000 |
| Anterior PV | Linear | -0.076 | 0.114 | 0.505 | 1.000 |
| Anterior PV | Quadratic | -0.262 | 1.070 | 0.806 | 1.000 |
| Right posterior horn | Intercept | 0.013 | 0.028 | 0.640 | 1.000 |
| Right posterior horn | Linear | -0.011 | 0.128 | 0.933 | 1.000 |
| Right posterior horn | Quadratic | -0.182 | 1.207 | 0.880 | 1.000 |
| Juxtacortical-deep occipital | Intercept | -0.023 | 0.028 | 0.412 | 1.000 |
| Juxtacortical-deep occipital | Linear | 0.097 | 0.117 | 0.406 | 1.000 |
| Juxtacortical-deep occipital | Quadratic | -0.904 | 1.107 | 0.414 | 1.000 |
| Temporal | Intercept | 0.026 | 0.027 | 0.321 | 1.000 |
| Temporal | Linear | -0.073 | 0.097 | 0.449 | 1.000 |

| WMH Measure | Effect | β | SE | p | pFDR |
|---|---|---|---|---|---|
| Temporal | Quadratic | -0.042 | 0.892 | 0.962 | 1.000 |
| Deep peri basal ganglia | Intercept | -0.085 | 0.028 | 0.002 | 0.132 |
| Deep peri basal ganglia | Linear | 0.296 | 0.116 | 0.011 | 0.624 |
| Deep peri basal ganglia | Quadratic | -2.479 | 1.083 | 0.022 | 1.000 |
| WMH volume | Intercept | -0.072 | 0.038 | 0.057 | 1.000 |
| WMH volume | Linear | 0.026 | 0.154 | 0.867 | 1.000 |
| WMH volume | Quadratic | -1.303 | 1.457 | 0.371 | 1.000 |

*Table 8: Associations of WMH spatial components with baseline and change in verbal memory function in LIFE. Standardized regression estimates are from the latent class mixed-effect models with WMH components and total WMH volume as predictors for intercept and change of memory, adjusting for age, sex and education. FDR-corrected p-values are shown. WMH: white matter hyperintensities, FDR: false-discovery rate, SE: standard error.*

| WMH Measure | Model | β | SE | p | pFDR |
|---|---|---|---|---|---|
| Fronto-Parietal | Intercept | -0.156 | 0.038 | 5.00e-05 | 0.004 |
| Fronto-Parietal | Change | -0.075 | 0.054 | 0.167 | 1.000 |
| Left posterior horn | Intercept | -0.116 | 0.038 | 0.00194 | 0.147 |
| Left posterior horn | Change | -0.067 | 0.051 | 0.194 | 1.000 |
| Anterior PV | Intercept | -0.112 | 0.038 | 0.00293 | 0.220 |
| Anterior PV | Change | -0.069 | 0.052 | 0.182 | 1.000 |
| Right posterior horn | Intercept | -0.136 | 0.038 | 3.00e-04 | 0.025 |
| Right posterior horn | Change | -0.068 | 0.051 | 0.186 | 1.000 |
| Juxtacortical-deep occipital | Intercept | -0.025 | 0.036 | 0.494 | 1.000 |

| WMH Measure | Model | β | SE | p | pFDR |
|---|---|---|---|---|---|
| Juxtacortical-deep occipital | Change | -0.081 | 0.051 | 0.113 | 1.000 |
| Temporal | Intercept | -0.103 | 0.038 | 0.00647 | 0.466 |
| Temporal | Change | -0.111 | 0.053 | 0.0378 | 1.000 |
| Deep peri basal ganglia | Intercept | -0.097 | 0.036 | 0.00753 | 0.527 |
| Deep peri basal ganglia | Change | 0.015 | 0.058 | 0.793 | 1.000 |
| WMH volume | Intercept | -0.150 | 0.039 | 1.10e-04 | 0.009 |
| WMH volume | Change | -0.100 | 0.054 | 0.0635 | 1.000 |

### 9 Dementia and stroke results

*Table 9: Associations of WMH spatial components with all-cause dementia incidence in 3C-Dijon from Cox proportional hazard models adjusting for age, sex, education and TIV.*

| WMH measure | HR | 2.5% CI | 97.5% CI | p | pFDR |
|---|---|---|---|---|---|
| Fronto-Parietal | 1.11 | 0.94 | 1.33 | 0.224 | 0.90 |
| Left posterior horn | 0.86 | 0.72 | 1.02 | 0.088 | 0.44 |
| Anterior PV | 1.27 | 1.05 | 1.53 | 0.012 | 0.08 |
| Right posterior horn | 1.10 | 0.92 | 1.33 | 0.293 | 0.90 |
| Juxtacortical-deep occipital | 1.22 | 1.00 | 1.47 | 0.045 | 0.27 |
| Temporal | 0.97 | 0.81 | 1.16 | 0.716 | 1.00 |
| Deep peri basal ganglia | 0.99 | 0.83 | 1.18 | 0.910 | 1.00 |
| WMH volume | 1.37 | 1.08 | 1.74 | 0.010 | 0.08 |

*Table 10: Associations of WMH spatial components with probable or possible AD dementia incidence in 3C-Dijon from Cox proportional hazard models adjusting for age, sex, education and TIV.*

| WMH Measure | HR | 2.5% CI | 97.5% CI | P | pFDR |
|---|---|---|---|---|---|
| Fronto-Parietal | 1.11 | 0.91 | 1.35 | 0.32 | 0.78 |
| Left posterior horn | 0.87 | 0.70 | 1.07 | 0.18 | 0.78 |
| Anterior PV | 1.20 | 0.97 | 1.49 | 0.10 | 0.78 |
| Right posterior horn | 1.10 | 0.88 | 1.36 | 0.40 | 0.78 |
| Juxtacortical-deep occipital | 1.21 | 0.97 | 1.51 | 0.10 | 0.78 |
| Temporal | 0.86 | 0.70 | 1.05 | 0.14 | 0.78 |
| Deep peri basal ganglia | 0.89 | 0.72 | 1.09 | 0.25 | 0.78 |
| WMH volume | 1.25 | 0.95 | 1.65 | 0.11 | 0.78 |

*Table 11: Associations of WMH spatial components with any stroke incidence in 3C-Dijon (N=60) from Cox proportional hazard models adjusting for age, sex and TIV.*

| WMH Measure | HR | 2.5% CI | 97.5% CI | P | pFDR |
|---|---|---|---|---|---|
| Fronto-Parietal | 1.39 | 1.09 | 1.78 | 0.0073 | 0.12 |
| Left posterior horn | 1.03 | 0.80 | 1.32 | 0.81 | 1 |
| Anterior PV | 1.67 | 1.28 | 2.18 | 1.50e-04 | 0.0036 |
| Right posterior horn | 1.20 | 0.93 | 1.54 | 0.17 | 1 |
| Juxtacortical-deep occipital | 1.32 | 1.02 | 1.72 | 0.038 | 0.56 |
| Temporal | 1.22 | 0.95 | 1.56 | 0.11 | 1 |
| Deep peri basal ganglia | 1.16 | 0.91 | 1.46 | 0.23 | 1 |
| WMH volume | 1.80 | 1.32 | 2.45 | 2.10e-04 | 0.0048 |

*Table 12: Associations of WMH spatial components with ischemic stroke incidence in 3C-Dijon (N=50) from Cox proportional hazard models adjusting for age, sex, education and TIV.*

| WMH Measure | HR | 2.5% CI | 97.5% CI | P | pFDR |
|---|---|---|---|---|---|
| Fronto-Parietal | 1.25 | 0.94 | 1.64 | 0.12 | 1 |
| Left posterior horn | 1.03 | 0.77 | 1.36 | 0.86 | 1 |
| Anterior PV | 1.54 | 1.14 | 2.08 | 0.004 | 0.076 |
| Right posterior horn | 1.26 | 0.94 | 1.68 | 0.12 | 1 |
| Juxtacortical-deep occipital | 1.07 | 0.82 | 1.41 | 0.62 | 1 |
| Temporal | 1.21 | 0.92 | 1.61 | 0.18 | 1 |
| Deep peri basal ganglia | 0.96 | 0.73 | 1.26 | 0.76 | 1 |
| WMH volume | 1.36 | 1.04 | 1.78 | 0.023 | 0.36 |

*Table 13: Associations of WMH spatial components with hemorrhagic stroke incidence in 3C-Dijon (N=10) from Cox proportional hazard models adjusting for age, sex, education and TIV.*

| WMH Measure | HR | 2.5% CI | 97.5% CI | p | pFDR |
|---|---|---|---|---|---|
| Fronto-Parietal | 1.88 | 1.02 | 3.45 | 0.044 | 0.62 |
| Left posterior horn | 1.06 | 0.56 | 2.00 | 0.86 | 1 |
| Anterior PV | 3.39 | 1.58 | 7.29 | 0.002 | 0.04 |
| Right posterior horn | 1.32 | 0.68 | 2.57 | 0.41 | 1 |
| Juxtacortical-deep occipital | 3.69 | 1.51 | 9.05 | 0.004 | 0.076 |
| Temporal | 1.42 | 0.77 | 2.62 | 0.27 | 1 |
| Deep peri basal ganglia | 2.39 | 1.43 | 3.99 | 0.001 | 0.021 |
| WMH volume | 2.51 | 1.46 | 4.32 | 8.91e-04 | 0.02 |
